# Distinct Neurodevelopmental Signatures of Sex-Specific Transcriptome-Based Polygenic Risk Scores for Depression

**DOI:** 10.1101/2025.07.31.25332511

**Authors:** Sarah S. Rashid, Kevan P. Clifford, Amy E. Miles, Fernanda C. Dos Santos, Etienne Sibille, Yuliya S. Nikolova

**Author notes:** **Corresponding author:** Yuliya S. Nikolova, PhD Independent Scientist, Centre for Addiction and Mental Health 250 College St Toronto, ON M5T 1R8 Canada.

## Abstract

Major depressive disorder (MDD) impacts females and males differently and post-mortem gene expression profiling reveals distinct transcriptomic signatures of the disorder in each sex. Using genes that are transcriptionally altered in MDD in both sexes, we recently developed a novel transcriptome-based polygenic risk score (tPRS), which had sex-specific associations with brain structure and depressive symptoms in both adolescents and adults. Identifying the neurodevelopmental signatures of genetically-induced shifts toward a depression-like brain transcriptome in each sex during a crucial stage, when sex differences in MDD vulnerability initially manifest, could provide useful information about the developmental pathways of early MDD risk. Leveraging sex-specific MDD gene expression data, we sought to develop female- and male-specific tPRSs (tPRS-F and tPRS-M, respectively) and evaluate their impact on regional cortical thickness, cortical surface area, subcortical volume, and depressive symptoms at baseline and 2-year follow-up in a developmental sample of 5002 adolescents (46.6% female, aged 8.9-11.0). In males, tPRS-M was associated with higher depressive symptoms at both timepoints and thicker left posterior cingulate at follow-up. In females, tPRS-F was associated with lower volumes of the right accumbens area, right caudate, and bilateral hippocampi at follow-up. Subcortical volumes of the right caudate and right hippocampus further mediated an indirect effect of tPRS-F on depressive symptoms. For each sex-specific PRS, no effects emerged in the opposite sex. Our findings suggest that sex-specific depression-like shifts in gene expression may contribute to unique vulnerability phenotypes for future MDD risk via distinct mechanisms in each sex.

## Introduction

Major depressive disorder (MDD), or depression, is a heritable psychiatric disorder characterized by mood impairment, anhedonia, and heterogeneous physiological features (1), often comorbid with anxiety (2–4). MDD is twice as prevalent in women relative to men (5). The disorder also impacts men and women differently, e.g., women report a greater number of symptoms, more severe illness, and different symptom profiles than men (6–9). Women may also be more susceptible to recurrence than men (10). Although this disparity is partially due to gender-related psychosocial differences (11–14), biological sex plays a critical role in the etiology of MDD (11,14).

Consistent with this notion, post-mortem transcriptomic studies show distinct sex-specific molecular signatures of MDD. In corticolimbic brain regions implicated in MDD – the dorsolateral prefrontal cortex (DLPFC), anterior cingulate cortex (ACC), and amygdala, a sex-stratified meta-analysis found opposite gene expression changes in each sex (15). Specifically, there was significant overlap of genes changed in opposite directions in the DLPFC and ACC of males and females with MDD. Males with MDD had downregulated expression of synapse-related genes and upregulated expression of oligodendrocyte- and microglia-related genes, whereas women with MDD exhibited the opposite patterns in these pathways (15). In another study, using RNA-sequencing (RNA-seq) to examine differential gene expression across six brain regions – the ventromedial PFC (VMPFC), OFC, DLPFC, anterior insula, nucleus accumbens, and ventral subiculum, significant transcriptional changes were identified across each of these brain regions in males and females with MDD as compared to healthy controls, with little overlap between the two sexes (16).

Despite the identification of MDD-associated transcriptomic changes through gene expression profiling in the post-mortem brain, there remains no direct method of evaluating the link between these molecular patterns and *in vivo* brain structural and functional changes or clinically relevant symptoms in living individuals. To better understand the functional impact of MDD-related transcriptomic alterations on the living brain, our group developed a transcriptome-based polygenic risk score (tPRS) and assessed its relationship with brain structure and function and depressive symptoms (17–19). We used *cis*-expression quantitative trait loci (*cis*-eQTLs), which are single nucleotide polymorphisms (SNPs) that affect variation in gene expression levels, acting as non-invasive proxies to index the differentially expressed MetaA-MDD genes in *in vivo* samples (17). Although this tPRS was based on non-sex-specific transcriptomic signatures, it was associated with sex-specific patterns of neural activity during face perception, MDD risk and symptom severity, as well as individual differences in cortical gyrification, subcortical volume (SCV), and cortical thickness (CT) in young adults (17–18) and adolescents (19). These phenotypic signatures partially converged with, but were notably distinct from, those associated with polygenic risk scores (PRS) computed based on conventional methods using genome-wide association studies (GWAS) summary statistics (MDD-PRS; 20), suggesting tPRS may capture complementary pathways of risk not reflected in other methods.

Although these phenotypic effects emerged in young adults, skewed sex ratios in depression risk first start to emerge in adolescence (21). The period of adolescence is a crucial stage for the maturation of the brain and the emergence of psychiatric disorders (22), during which sex hormones remodel and activate neural circuits (23). Sex differences stemming from biological developmental processes may play a more significant role in the developmental origins of MDD, as opposed to differences between men and women arising in adulthood, which may be attributable to environmental effects and more strongly reflect gender-based factors (24). Delineating the neurodevelopmental signatures of genetically driven shifts toward a more depression-like brain transcriptome separately in each sex, during a critical period when sex differences in MDD vulnerability first emerge, may therefore yield potentially actionable insight into developmental pathways of early MDD risk.

Given the heterogeneity of MDD (25), along with well-documented sex differences in MDD biology, as well as inter-regional differences in gene expression patterns (26), tPRS-like approaches can be improved by developing sex- and brain-region-specific scores, based on transcriptomic signatures of specific MDD subtypes. Recurrent MDD (rMDD) represents a more severe subtype of MDD that may have a stronger genetic component (27–29) and tends to be more common in individuals assigned female at birth (10,30,31). Thus, we sought to develop two tPRSs – a female-specific tPRS (tPRS-F) and a male-specific tPRS (tPRS-M) – based on gene expression in the DLPFC uniquely associated with rMDD in each sex, and to evaluate their impact on brain structure and early depression vulnerability in a developmental sample drawn from the Adolescent Brain Cognitive Development ^SM^ Study (ABCD Study^®^; 32).

## Methods

### 3.1 Participants

The sample consisted of participants from the ABCD Study^®^ (32). For the current analyses, the sample was restricted to 5151 participants of European ancestry to match the ethnic background of the post-mortem cohorts used to develop the tPRSs (15,33). Quality control (QC) of genetic data was conducted as described by Wainberg and colleagues (34) and 10 genetic principal components (PCs) were computed to account for residual population substructure in all analyses.

To preclude confounding effects of family structure in our sample, we identified pairs of relatives using proportional identity by descent (PIHAT). When we found pairs of individuals with PIHAT>0.2, we removed one individual from each pair from the analyses. In cases where one individual was part of multiple relative pairs, we excluded this particular individual from the analyses where applicable. In total, across analyses we excluded 147 individuals from 999 relative pairs using this approach (PIHAT=0.20–0.34). After this procedure, the sample was reduced to 5002 participants (2664 males, 2338 females; aged 8.9–11.0 years (M=9.9±0.6 years)) at baseline (Time 1 or T1). Of these, tabulated neuroimaging data was available for 4977 participants (2657 males, 2320 females; aged 8.9–11.0 years). At 2-year follow-up (Time 2 or T2), Child Behaviour Checklist (CBCL) data was available for 4569 participants (2443 males, 2126 females; aged 10.6–13.8 years (M=12.0±0.7 years)) and neuroimaging data was available for 3571 participants (1979 males, 1592 females; aged 10.6–13.7 years). At both time points, CBCL data was available for all 4569 of those participants but neuroimaging data was available for 3560 participants (1976 males, 1584 females).

### 3.2 Calculation of Sex-Specific Transcriptome-Based Polygenic Risk Scores (tPRS-F and tPRS-M)

ABCD procedures for DNA extraction and genotyping are described in detail by Uban et al (36). Imputation and QC of all genotype data from European-ancestry participants was conducted as previously described (35). Female- and male-specific gene lists from the DLPFC were derived from a post-mortem case-control comparison report (15). As described by Seney et al (15), post-mortem brain samples were obtained from the MD2 patient cohort, which consisted of 28 male and 30 female subjects, of which 50% were MDD cases. All cases were diagnosed with rMDD. Healthy controls (HCs) were unaffected by MDD and were matched to cases by age, sex, and race.

As previously described (15), RNA expression in tissue from the DLPFC was analyzed using the Affymetrix Human Genome U133 Plus 2.0 Array platform, using 53,596 oligonucleotide probes (or probe sets). We removed single probe sets that were matched to multiple genes and probe sets that were not matched to any gene symbol from our analyses. When multiple probes were matched to an identical gene symbol, we selected the probe that presented the greatest average effect size to represent the target gene, leaving us with a total of 19,944 genes. Using the Bioconductor BiomaRt R package (36), we were able to match 17,959 of these genes to Ensembl IDs.

We indexed the expression of these genes in DLPFC brain tissue from the CommonMind Consortium (CMC), an independent post-mortem brain tissue cohort, which served as a reference transcriptome (37). Using MetaXcan, an approach which uses elastic net regression to predict gene expression based on individual *cis*-eQTL SNPs (38), we effectively imputed the predicted relative cortical expression of these genes at the individual level for ABCD participants based on SNPs genotyped in peripheral tissue.

A DLPFC-specific prediction model (DLPFC_newMetax.db) was used to determine individual *cis*-eQTL SNP contributions. The expression levels of 9,345 genes were imputed for ABCD participants, which included 7,028 of the 17,959 genes for which we had effect sizes and Ensembl IDs. Once imputed, we weighted each gene by sex-specific average effect size (representing the expression levels in the MDD sample vs HCs) and direction of effect from the original post-mortem meta-analysis (15), and then combined them into tPRS-F or tPRS-M for all participants, regardless of sex. Hence, each tPRS was computed as the sum of the weighted expression values of the 7,028 imputed genes, with a higher tPRS-F or tPRS-M respectively representing a more female- or male-pattern depression-like transcriptome.

### 3.3 Neuroimaging Data

ABCD guidelines for MRI acquisition and preprocessing are described in detail by Hagler et al (39). Tabulated neuroimaging data was downloaded from the ABCD data repository. FreeSurfer-derived outputs were selected for subcortical and cortical hemisphere-specific regions-of-interest (ROIs) using automatic subcortical segmentation and Desikan-Killiany (DK) atlas parcellation (40), respectively. The following brain morphology metrics were used in our analyses: (1) SCV in each of the following subcortical ROIs: thalamus, caudate, putamen, pallidum, hippocampus, amygdala, accumbens area, and (2) CT and (3) cortical surface area (CSA) in each of the 34 DK atlas-based cortical ROIs. Due to the presence of extreme outliers, the data at each time point underwent a 90% winsorization, i.e., it was transformed by setting all values below the 5th percentile to the 5th percentile and all values above the 95th percentile to the 95th percentile. This winsorization approach was chosen in preference over removing all values more than 3 standard deviations away from the mean across all structural metrics, as the latter approach would remove 1540 participants (30.94% of our sample) from our neuroimaging analyses.

### 3.4 Depression Vulnerability Data

Scored data from the parent-reported Child Behavior Checklist (CBCL/6-18) was downloaded from the ABCD data repository. Its collection is described in detail by Barch and colleagues (41). CBCL data was collected every 6 months in the ABCD sample. We selected raw scores on the Anxious/Depressed Syndrome Scale (Anx/Dep) and Withdrawn/Depressed Syndrome Scale (With/Dep) for the same two time points that the neuroimaging data was available for (i.e., baseline and 2-year follow-up). We used raw scores instead of t-scores to account for the full range of variation in the sample, as recommended by the developers of the CBCL (42). Although the data distributions for both CBCL subscales were positively skewed, to preserve interpretability of the final results, we did not apply any transformation to the subscale scores. Importantly, our sample size is sufficiently large to yield reliable results even in the presence of skewed dependent variables (43). Furthermore, we decided against winsorization of these variables as skewed distributions of depressive symptom scales are typical for population-representative samples and winsorization would likely result in the loss of meaningful variability in the high end of the observed score range.

### 3.5 Statistical Analyses

#### 3.5.1 Cross-sectional Analyses

Linear mixed-effects models (LMMs) were used to test the effects of each tPRS on four sets of variables: (1) SCV, (2) CT, (3) CSA, and (4) CBCL scores. In our main models, we tested the effect of one of the sex-specific scores (either tPRS-F or tPRS-M) without including the opposite score (Model 1). This was done independently at T1 and T2. For variables with a significant association with either tPRS, we then re-ran the LMMs while including the opposite sex-specific score as a covariate (Model 2) to confirm the sex-specificity of the effect. We also ran additional LMMs including the original tPRS (non-sex-specific) and an MDD-PRS based on the PGC GWAS findings (20) (calculation of this score is described previously (17)), in addition to the covariates included in Model 2 (Model 3) to confirm that any sex-specific PRS effects explained phenotypic variance above and beyond that accounted for by these non-sex-specific PRSs.

Age and PCs were modeled as fixed effects and study site was modeled with a random intercept in all analyses. Estimated total intracranial volume (eTIV) was modeled as an additional fixed effect for SCV and CSA analyses. The false discovery rate (FDR) method of multiple comparison correction was applied within each of the four families of analyses, where significance was set to pFDR<0.05. In addition, we sought to identify more subtle, distributed patterns of cortical changes in regions that were significant at the uncorrected level (p<0.05). All analyses were performed in R version 4.2.2. When there was a significant effect of the tPRS on regional brain morphology in a particular sex, the effect of brain morphology in tPRS-associated regions on depressive symptoms was tested in the respective sex. If a significant association was found, then a causal mediation analysis was conducted using nonparametric bootstrapping to test the relationship among the tPRS, covariate-adjusted region-specific brain morphology, and depressive symptoms.

#### 3.5.2 Longitudinal Analyses

For participants with data available for both time points, we ran two longitudinal LMMs for the outcome variables that had a significant association with either tPRS at either time point. To test if the effect of tPRS was time-dependent, we performed a tPRS-by-time interaction model. If a significant interaction was found, we ran an additional “delta (Δ) model” where we analyzed the association between the tPRS and the change in the variable over time (value at T2 minus value at T1). If no significant interaction was found, we also tested if the association with tPRS was significant regardless of time, i.e., when considering data across the two time points. The fixed effects remained the same as those in previous models. For the random effects, in addition to study site with a random intercept, subject ID was included with time as a random slope. Since these analyses were conducted *post hoc* only for the variables showing significant cross-sectional associations with either tPRS, significance was set to p<0.05. To account for the discrepancy in sample size between time points, if an effect was significant in our longitudinal model, we ran sensitivity analyses to test the effect of tPRS at T1 and T2, using only participants with data available for both time points.

## 4 Results

### 4.1 Sample Characteristics

The two sex-specific tPRSs were negatively correlated (r=-0.115, p<2.2e-16), in line with the opposite molecular signatures observed between males and females in the original post-mortem case-control comparison (15). Intriguingly, each of the two scores was also positively associated with our previously described (19) non-sex-specific tPRS (tPRS-M: r=0.127, p<2.2e-16; tPRS-F: r=0.143, p<2.2e-16).

### 4.2 Effects of tPRS-M and tPRS-F on Depressive Symptoms

In females, tPRS-F was not directly associated with depressive symptoms (all p>0.1). In contrast, in males, tPRS-M was associated with higher depressive symptoms indexed by the With/Dep Scale at T1 (t=2.566, pFDR=.010; **Figure 1a**) and by the Anx/Dep Scale at both T1 (t=3.192, pFDR=.002; **Figure 1b**) and T2 (t=2.739, pFDR=.012; **Figure 1c**). Sensitivity analyses showed that in the subsample of male participants with data available for both time points, the effect of tPRS-M on Anx/Dep at both time points was significant (T1: t=2.970, p=.003; T2: t=2.856, p=.004), suggesting none of the effects were driven by biased longitudinal sampling. Neither sex-specific score was associated with depressive symptoms in the opposite sex at either time point (**Supplementary Tables S1 and S2**). The significance of all effects remained unchanged in Model 2, where the tPRS corresponding to the opposite sex was included as a covariate, as well as in Model 3, where tPRS and MDD-PRS were included as additional covariates.

**Figure 1.**
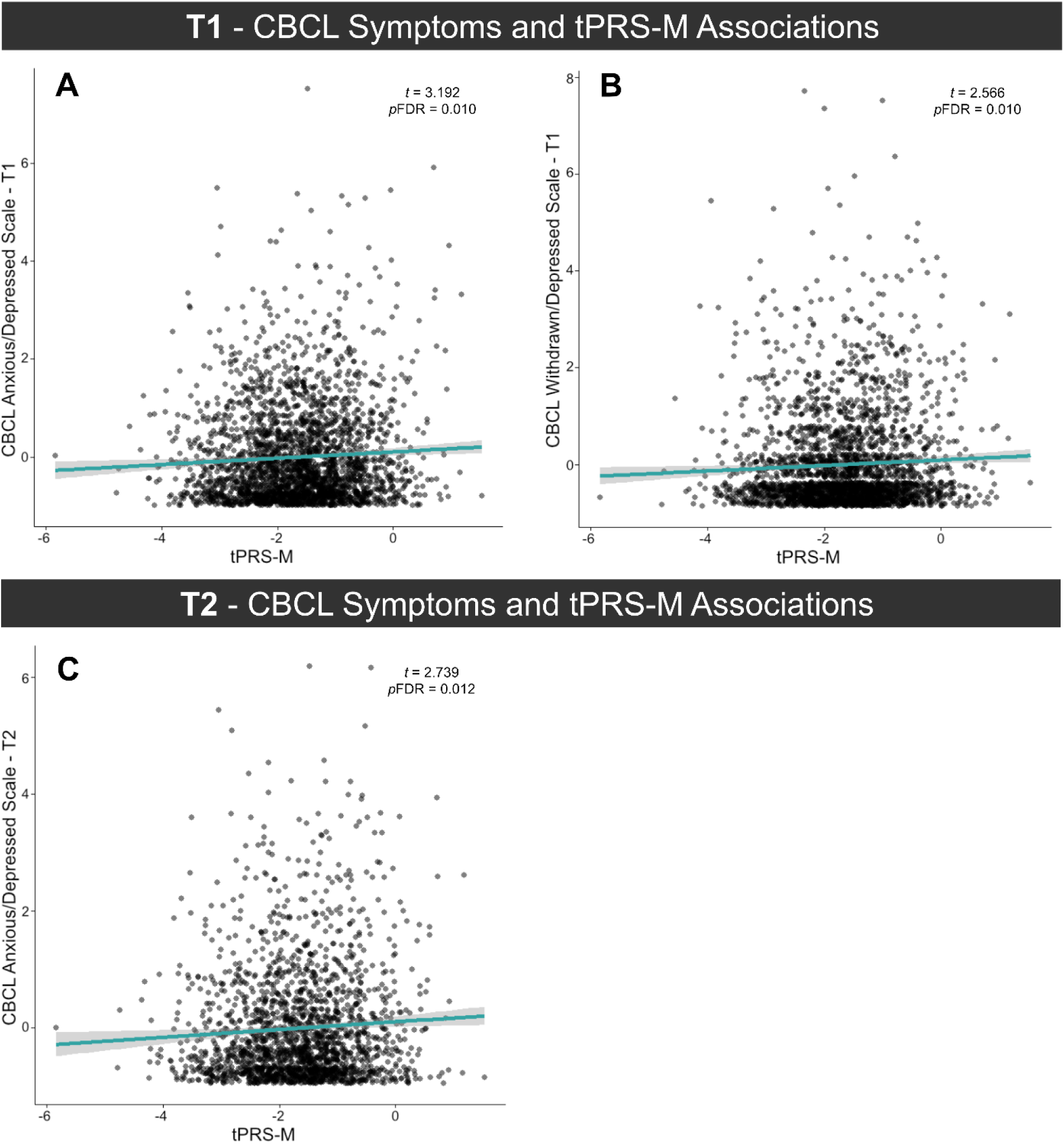
tPRS-M and depressed symptoms in males at T1 and T2. A significant association occurred at T1 between tPRS-M and higher **(a)** withdrawn/depressed symptoms, while similar associations with anxious/depressed symptoms occurred at both T1 **(b)** and at T2 **(c).** Age, 10 genetic principle components, and study site were regressed out of the CBCL scale measures.

### 4.3 Effects on Brain Structure

#### 4.3.1. Effects of tPRS-F on brain structure in females

No significant associations with cortical brain structure surviving FDR correction emerged at T1 for tPRS-F (**Supplementary Tables S3 and S4**). However, a distributed pattern of nominal associations occurred between tPRS-F and lower CT in frontotemporal regions at T1, and with greater CT in cingulate regions at T2. Conversely, nominally significant associations between tPRS-F and lower CSA were present in the left opercularis at T1 (t=-2.728, p=0.006) and left precentral (t=-2.985, p=0.02) at T2. All tPRS-F associations at T1 and T2 with CT are depicted in **Supplementary Figure S1**, and with CSA in **Supplementary Figure S2**.

At T2, higher tPRS-F was significantly associated with smaller subcortical volume in the left hippocampus (HPC.L; t=-2.985, pFDR=.038), right hippocampus (HPC.R; t=-2.715, pFDR=.038), right caudate (Cd.R; t=-2.599, pFDR=.038), and right accumbens area (Ac.R; t=-2.546, pFDR=.038) (**Figure 2a-b**). Lower volumes of the right hippocampus and right accumbens area were in turn associated with higher symptoms measured by the CBCL With/Dep scale (HPC.R: t=-2.039, p=.042; Ac.R: t=-3.038, p=.002) at T2. Hippocampal volume mediated an indirect relationship between higher tPRS-F and depressive symptoms (HPC.R: estimated average causal mediation effect (ACME_est)=.0036, 95% CI: 0.0004-0.0076, p=.012), while the mediation effect fell just short of significance for the accumbens area (Ac.R: ACME_est=0.0032, 95% CI: -0.00005-0.0079, p=.056; **Figure 2c-d**).

**Figure 2.**
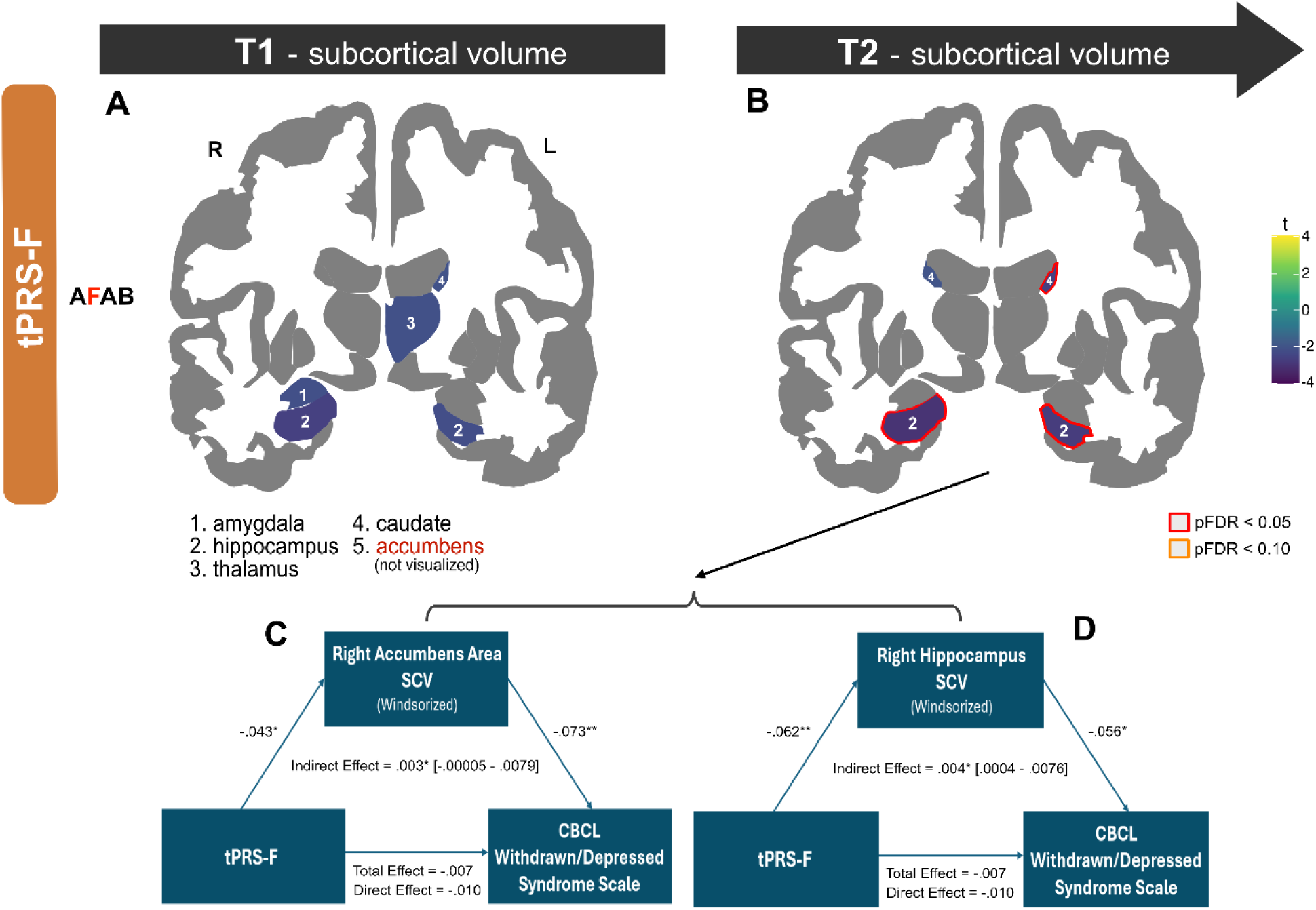
tPRS-F and regional subcortical volume in females at T1 and T2, and associated regional mediations. Nominally and pFDR significant lower volume in subcortical structures are depicted at T1 **(a)** and T2 **(b)**. Indirect effects of tPRS-F on withdrawn/depressed symptoms were significantly mediated by the right accumbens **(c)** and right hippocampus **(d)**. 10 genetic principle components, eTIV, and study site were regressed out of the subcortical volume measure in each plot. *p < 0.1 (’); p < 0.05 (*); p < 0.01 (**)*.

We also detected a significant tPRS-F-by-time interaction effect for the left hippocampus (t=-2.247, p=.025), where in addition to being associated with a smaller hippocampal volume at T2, tPRS-F was also associated with smaller increase, or in some participants greater decrease, in hippocampal volume between the two time points (t=-2.108, p=.035).

No significant time-by-tPRS-F interactions emerged for the right hippocampus, caudate, and accumbens area (all p>0.05; **Supplementary Table S5**), suggesting the association between tPRS-F and volume in these regions was consistent between time points, even if it did not survive FDR correction at T1. Indeed, the association of tPRS-F over time was significant for the right hippocampus (t=-2.504 p=.012) and caudate (t=-2.434, p=.015) in a longitudinal LMM. Sensitivity analyses showed consistent results in the subsample of female participants with data available for both time points (T1: HPC.R: t=-2.276, p=.023; Cd.R: t=-2.932, p=.003; T2: HPC.R: t=-2.780, p=.006; Cd.R: t=-2.724, p=.007).

All significant effects remained significant in Models 2 and 3. As expected, no significant effects on brain structure were found for tPRS-F in males.

#### 4.3.2. Effects of tPRS-M on brain structure in males

Similarly to tPRS-F, no significant associations with cortical brain structure surviving FDR correction emerged at T1 for tPRS-M (**Supplementary Tables S6 and S7**). However, there was a distributed pattern of nominally significant associations between tPRS-M and greater CT in frontal and occipital regions (**Figure 3a**) and greater CSA in temporal regions (**Supplementary Figure S3**).

**Figure 3.**
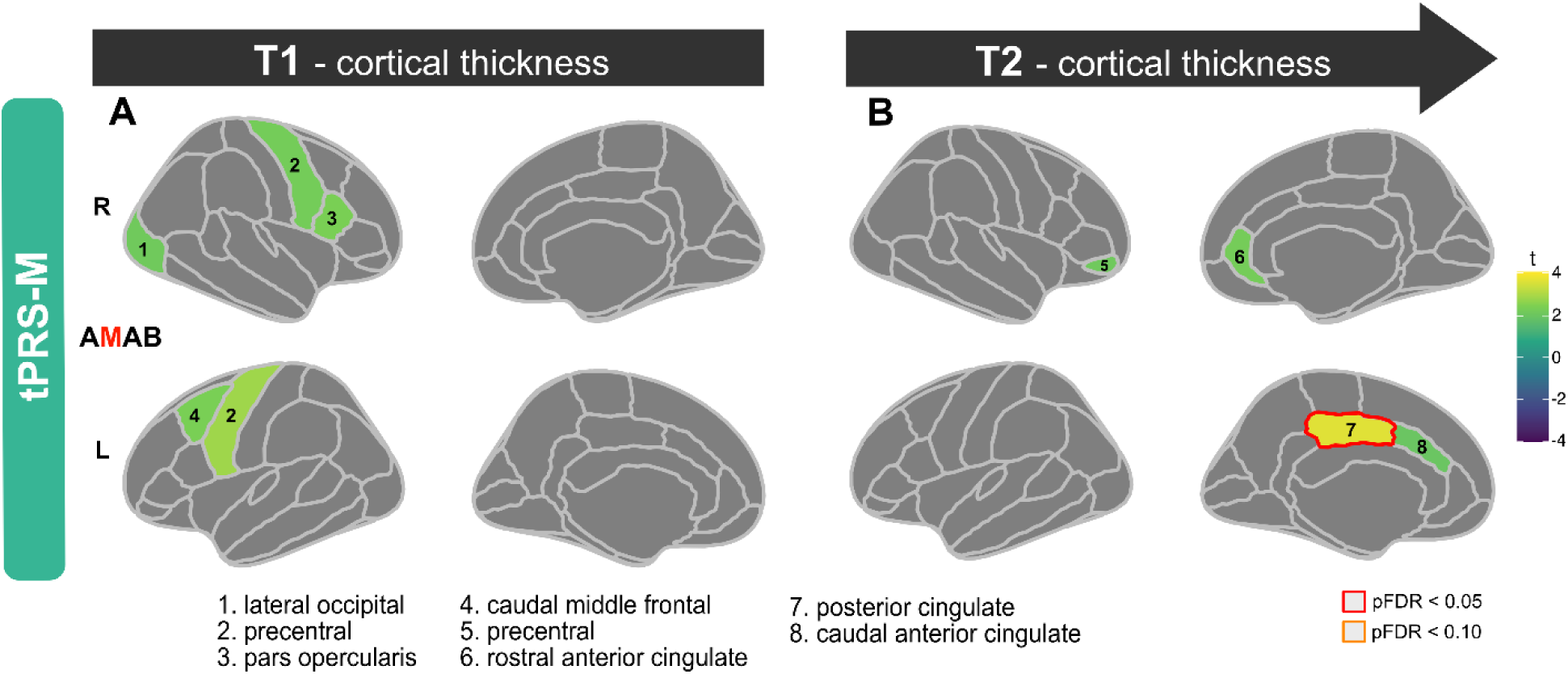
tPRS-M cortical thickness effects in males at T1 and T2. Associations between tPRS-M and cortical thickness in males are depicted at T1 **(a)** and T2 **(b)**. At T2, the right posterior cingulate demonstrated pFDR significant increased cortical thickness. 10 genetic principle components, and study site were regressed out.

In contrast, at T2 higher tPRS-M was robustly associated with greater CT of the left posterior cingulate after FDR-correction (PC.L; t=3.626, pFDR<.001; **Figure 3b**). This effect remained significant in Model 2, where tPRS-F was included as a covariate, as well as in Model 3, where tPRS and MDD-PRS were included as additional covariates. However, this region did not have a significant association with symptoms measured by the CBCL (With/Dep: t=-0.058, p=.953; Anx/Dep: t=1.406, p=.160). No significant tPRS-M-by-time interaction emerged for the left posterior cingulate (t=0.735, p=.462), suggesting the effect of tPRS-M was consistent across the two time points, despite not passing FDR correction at T1. Indeed, the association of tPRS-M over time was significant for this region at an uncorrected level (t=3.689, p<.001). This association remained significant in Models 2 and 3. Sensitivity analyses revealed that in male participants with data available for both time points, the effect of tPRS-M on this region was significant at both T1 (t=3.136, p=.002) and T2 (t=3.641, p<.001).

As expected, no significant effects on brain structure surviving FDR correction were found for tPRS-M in females (**Supplementary Table 6**).

## Discussion

In this study, we developed novel female- and male-specific tPRSs (tPRS-F and tPRS-M, respectively) and identified their developmental neuroanatomical signatures and associations with depressive symptoms in each sex. In females only, tPRS-F was associated with brain structure alterations in four subcortical areas, and in males only, tPRS-M was associated with alterations in one cortical area as well as anxious/depressed and withdrawn/depressed symptoms. Neither of the scores had any associations with brain structure or depression vulnerability in the opposite sex. Moreover, our effects remained significant even after controlling for a broad GWAS-based MDD-PRS and a non-sex-specific tPRS. These findings suggest that sex-specific depression-like shifts in gene expression may increase risk for depression via distinct mechanisms in both sexes and contribute to a latent vulnerability phenotype for future MDD risk not captured by existing GWAS-based PRS approaches. Thus, divergent molecular changes associated with MDD in males and females may partially drive sex differences in MDD-related neuroanatomy and symptomatology.

In females, we detected an association between higher tPRS-F and lower volume in the bilateral hippocampi at T2. The hippocampus is a limbic structure responsible for receiving input from dorsal stream cortical areas which carry visual information from the primary visual cortex to the posterior parietal cortex, and plays a crucial role in learning and consolidating explicit memories from short-term memory to long-term memory storage in the cortex, as well as in the regulation of the HPA axis stress response (44–46). Reduced hippocampal volume in depression is well-documented (47,48) and has been linked to the neurotrophic hypothesis of depression, which posits that chronic hyperactivity of the HPA axis in MDD leads to brain atrophy (46). The rise in estrogens in female adolescents enhances the HPA axis stress response, whereas in male adolescents, androgens such as testosterone inhibit stress reactivity (49,50). Thus, this effect may be female-specific due to the upregulation of their stress response during this critical developmental period. Lower hippocampal volume from early to mid-adolescence has been linked to depression onset during adolescence, and may be a vulnerability factor for the development of depression later in life (51). Therefore, tPRS-F may contribute to an etiological mechanism for MDD through its effect on the hippocampus.

In females, higher tPRS-F was also associated with lower volume of two components of the striatum in the basal ganglia – the caudate and accumbens – which are part of the limbic-striato-pallidal-thalamo-cortical pathways involved in mood regulation (52). The caudate is part of the dorsal striatum and supports reward, motivation, learning, memory, and emotion (53). Studies have found lower caudate volume in adolescents (54) and women with MDD (55). The accumbens is part of the ventral striatum and plays a role in the regulation of emotional- and reward-related stimuli, as well as in motivation and learning (56). Thus, lower volume of this region may indicate reward-related dysfunction. Smaller nucleus accumbens volume across time in female adolescents, but not in males, has been associated with depression onset during adolescence (51). Research has also shown sex differences in the dopaminergic function of the posterior caudate nuclei and ventral striatum, with males exhibiting higher dopamine release than women in these regions (57). Dopamine system dysregulation is implicated in MDD, with lower levels of dopamine being linked with anhedonia and low motivation (58), which may help to explain the female-specificity of these effects.

Although we did not find significant direct effects of the tPRS-F on depressive symptoms, tPRS-F may indirectly influence symptoms through the mediating effect of lower volumes of the right hippocampus and accumbens area, which were associated with a higher tPRS-F in females, and further associated with more withdrawn/depressed symptoms. Studies have shown that children who were at risk for MDD based on familial history had lower right hippocampal and nucleus accumbens volume prior to onset (59–61), which suggests that these brain alterations may be involved in depression risk and onset.

In contrast to these subcortical findings in females, higher tPRS-M was associated with greater CT of the posterior cingulate in males. The posterior cingulate cortex (PCC), part of the default-mode network (DMN), is involved in topographic and topokinetic memory and visuospatial processing (62). MDD has been characterized by excessive activation of the DMN and research has shown that patients with MDD show hyperconnectivity between the PCC and other DMN regions, further linked to ruminative thoughts and pain (63,64). Our findings are consistent with meta-analytic research showing thicker left PCC in MDD patients relative to HCs (65). The association with a thicker posterior cingulate in males could be due to alterations in typical cortical maturation trajectories. In typical brain development, CT follows a straightforward linear reduction by age 8 (66), which has been proposed to reflect biological mechanisms such as synaptic pruning (67), myelination (68), and morphological changes that cause the neuropil (a synaptically dense region composed of glial and neuronal processes) to stretch (69). The relationship between a more depression-like cortical transcriptome and higher posterior cingulate thickness in young adolescents may point to abnormal neurodevelopment that delays cortical thinning and/or neuronal hyperactivation in this region. Although the mechanisms underlying increased CT in depression are not completely understood, it has been speculated that this phenotype may be due to activation of the immune-inflammatory response system and the compensatory immune-regulatory reflex system (65,70). These inflammatory pathways may lead to impaired pruning or increased density of the neuropil, which may lead to cortical thickening. Consistent with this, males with MDD showed an upregulation of genes expressed specifically in microglia, which are immune cells that play crucial roles in neuroinflammation (15). Thus, we speculate that the effect of tPRS-M on thicker posterior cingulate in males may be partially driven by these genes.

Examining broader, distributed patterns of nominal cortical associations with tPRS-F and tPRS-M revealed more subtle effects. Notably, alongside the pronounced posterior cingulate association in males at T2, both the rostral anterior cingulate and caudal anterior cingulate regions displayed similar, though nominal, greater CT, suggesting this effect may encompass broader cingulate cortex regions. Furthermore, these same cingulate regions were nominally associated with tPRS-F and greater CT in females at T2, possibly indicating altered cingulate cortex development shared between sexes, but more pronounced in males. Finally, while tPRSs were largely associated with patterns of greater CT, they were conversely associated with broad patterns of lower CSA, suggesting opposite directions of effects on these cortical phenotypes. Given the nominal nature of these effects, future work is needed to confirm these conjectures.

The significant tPRS-F-by-time interaction for the left hippocampus in females, whereby a higher tPRS-F was linked to less apparent growth of the left hippocampus over time, suggests that tPRS-F may be associated with altered neurodevelopmental trajectories in females – a finding which merits further investigation as ABCD participants are followed into early adulthood. Although we did not find significant tPRS-by-time interactions for the other significant regions, the effect sizes at T2 were still nominally larger and more significant than at T1. The overall strengthening of the sex-specific associations over time may be due to increasing levels of sex hormones during puberty, which exert different effects on brain development in the two sexes (71). Some sexually dimorphic features, such as cell density and connectivity, are primarily influenced by the presence or absence of the perinatal testosterone surge (organizational effects of hormones), whereas the maintenance of neuronal circuitry involved in reproductive behaviors requires adequate levels of circulating sex hormones during adulthood (activational effects) (71–73). As the levels of sex steroids start to rise and approach adult levels during puberty, hormone-dependent changes in neuronal circuitry mature gradually, which is evident in both cortical and subcortical regions (74,75).

The current results are important to consider in the context of a recent study (19), where we computed a non-sex-specific tPRS, comprising genes altered consistently across the sexes, and mapped its neurodevelopmental impact in the same ABCD sample. In this prior work, we found partially convergent associations between tPRS with a thicker left posterior cingulate across both sexes and timepoints, along with female-specific lower right pallidal volume at T1 and lower right hippocampal volume at both timepoints (19). Although we did not find any significant associations at T1 in the current study, two of the same regions were implicated at T2, i.e., posterior cingulate and hippocampus. Further, despite no associations with the pallidum in the current study, the pallidum is a component of the basal ganglia and part of the limbic-striato-pallidal-thalamo-cortical pathways, as are the caudate and accumbens, which had female-specific associations here. Similar to the effect of subcortical volume in the current study, pallidal volume mediated an effect of the score on withdrawn/depressed symptoms (19). Thus, both sex-specific and non-sex-specific scores consistently identified thicker posterior cingulate and smaller hippocampal or basal ganglia volume as reflective of transcriptomic risk for depression in youth.

Importantly, however, the neurostructural effects of the sex-specific scores at T2 remained when the non-sex-specific tPRS was controlled for, while the effect of tPRS was no longer significant when the sex-specific scores were included. This suggests that over time, PRSs capturing sex-specific transcriptomic risk for depression become more predictive of brain structure within each sex than a non-sex-specific score. Conversely, since the tPRS is non-sex-specific, its effects may be more clearly detectable at T1, when puberty-related sex-specific maturational processes are less pronounced across the sample. Notably, the fact that tPRS-F and tPRS-M are both positively correlated with the tPRS, while negatively correlated with each other, supports the existence of both sex-specific and non-sex-specific pathways of transcriptomic risk for depression. This highlights the value of modeling both types of processes in future work seeking to map trajectories of risk across adolescence and young adulthood.

This study had several limitations. Firstly, cortical gene expression could not be imputed for all genes in our gene lists. Future work using more advanced *cis*-eQTL predictive models as well as gene lists derived from larger post-mortem samples would allow further refinement of our scores. Another constraint is that our gene lists were produced through microarray analysis rather than RNA sequencing (RNA-Seq), which may be able to detect a larger number of differentially expressed genes (76). Future studies using RNA-Seq may help identify additional relevant genes and improve our tPRSs’ predictive power. Additionally, the current analyses included age as a covariate, but future work should also analyze the moderating effect of pubertal stages on tPRS-associated brain development and depressive symptoms. Further, the analyses were limited to participants of European ancestry to match the demographics of the postmortem samples used to derive the gene lists and *cis*-eQTL statistics (37) and improve statistical power to detect small effects. To enhance the tPRSs’ generalizability across ethnicities, future studies should include multi-ethnic samples and analytic strategies. Lastly, for consistency with post-mortem research to date, we focused our analysis on biological sex and did not consider the independent effects of gender-related variables. Future studies should examine the impact of our sex-specific tPRSs in gender-diverse populations and possibly seek to develop novel tPRSs that capture sex-and/or gender-related variability in the molecular signatures of MDD in a more comprehensive way.

Limitations aside, our findings demonstrate for the first time that genetically-driven shifts towards sex-specific depression-like gene expression patterns may have an early impact on neurostructural development and depressive symptoms in adolescence – a critical period for the emergence of sex differences in MDD vulnerability. Hence, our newly developed tPRSs may reflect etiological mechanisms leading to sex-specific patterns of atypical brain maturation that may contribute to the differential susceptibility to MDD in males and females, and inform the future development of personalized early intervention and prevention strategies.

## Data Availability

All data produced in the present work are contained in the manuscript

## Acknowledgements

YSN is supported by a Discovery Grant (RGPIN-2020-07131) from the Natural Sciences and Engineering Research Council of Canada (NSERC), a Koerner New Scientist Award, a Paul Garfinkel New Investigator Catalyst Award, and a womenmind™ Seed Grant administered by the Centre for Addiction and Mental Health Foundation.

Data used in the preparation of this article were obtained from the Adolescent Brain Cognitive Development^SM^ (ABCD) Study (https://abcdstudy.org), held in the NIMH Data Archive (NDA). This is a multisite, longitudinal study designed to recruit more than 10,000 children age 9–10 and follow them over 10 years into early adulthood. The ABCD Study® is supported by the National Institutes of Health and additional federal partners under award numbers U01DA041048, U01DA050989, U01DA051016, U01DA041022, U01DA051018, U01DA051037, U01DA050987, U01DA041174, U01DA041106, U01DA041117, U01DA041028, U01DA041134, U01DA050988, U01DA051039, U01DA041156, U01DA041025, U01DA041120, U01DA051038, U01DA041148, U01DA041093, U01DA041089, U24DA041123, U24DA041147. A full list of supporters is available at https://abcdstudy.org/federal-partners.html. A listing of participating sites and a complete listing of the study investigators can be found at https://abcdstudy.org/consortium_members/. ABCD consortium investigators designed and implemented the study and/or provided data but did not necessarily participate in the analysis or writing of this report. This manuscript reflects the views of the authors and may not reflect the opinions or views of the NIH or ABCD consortium investigators.

## Supplementary Figures

**Supplementary Figure 1.**
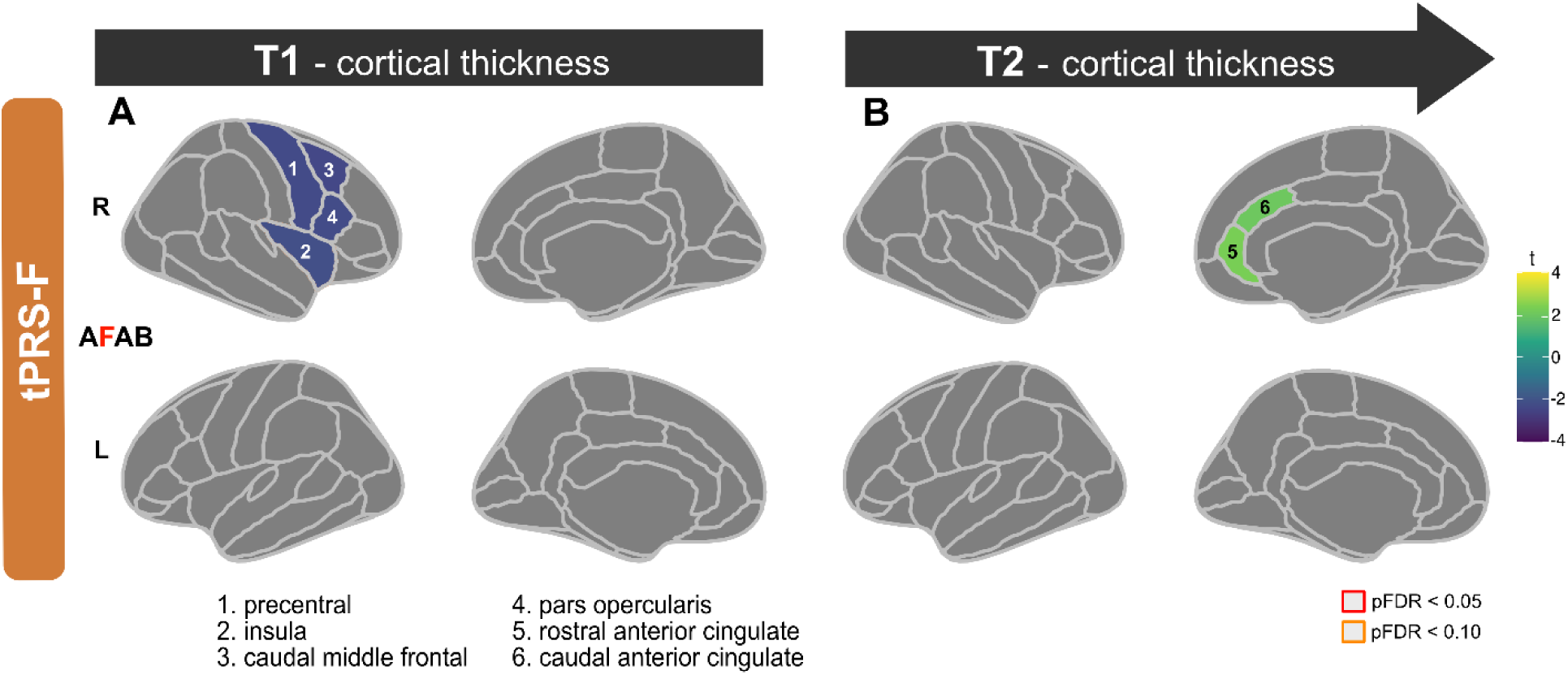
tPRS-F cortical thickness effects in females at T1 and T2. Associations between tPRS-F and cortical thickness in females are depicted at T1 **(a)** and T2 **(b)**. 10 genetic principle components and study site were regressed out.

**Supplementary Figure 2.**
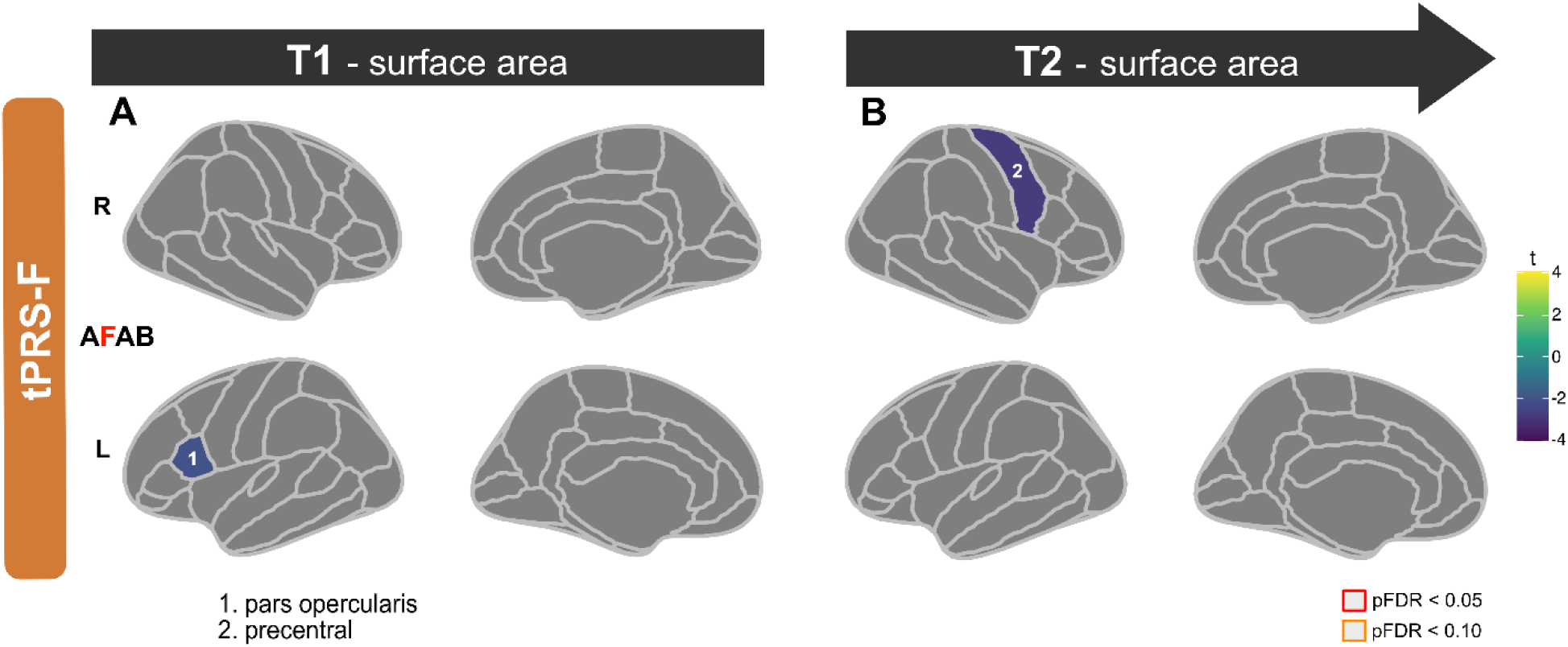
tPRS-F cortical surface area effects in females at T1 and T2. Associations between tPRS-F and cortical surface area in females are depicted at T1 **(a)** and T2 **(b)**. 10 genetic principle components, eTIV, and study site were regressed out.

**Supplementary Figure 3.**
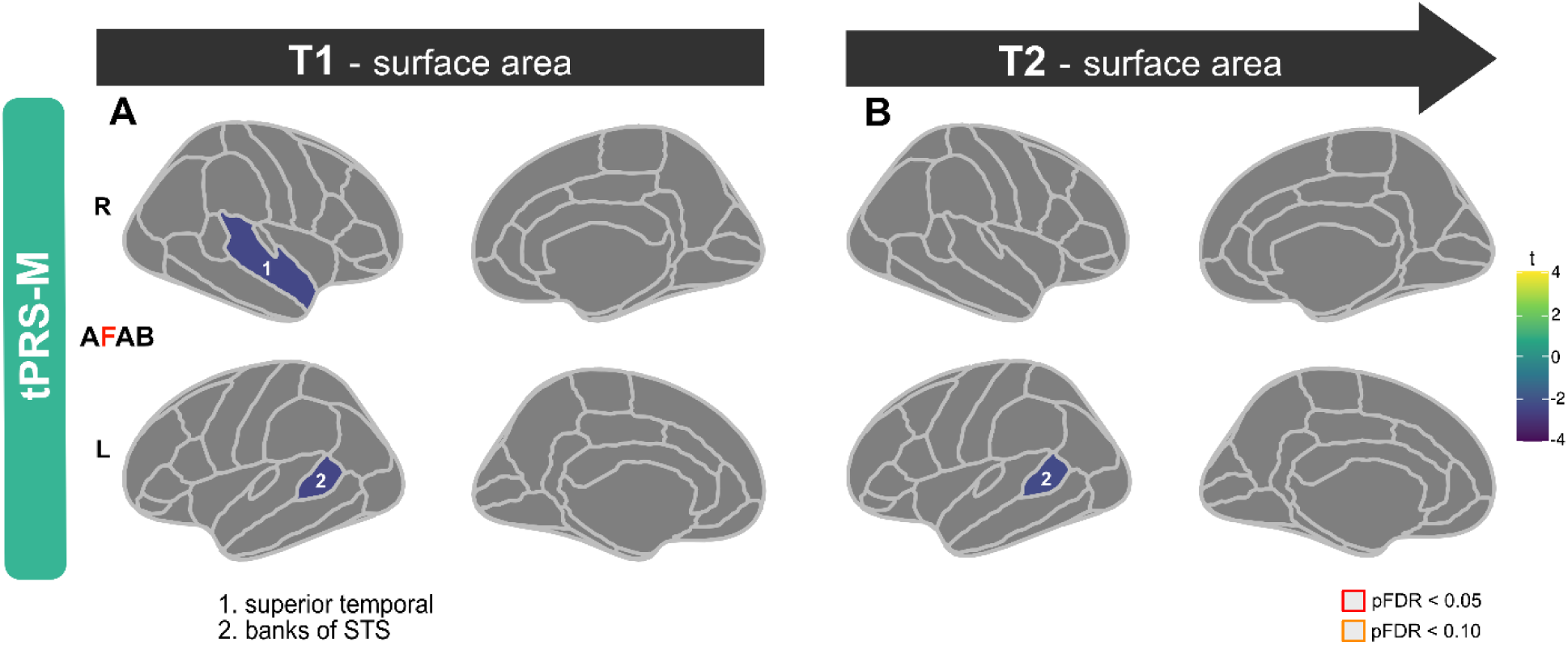
tPRS-M cortical surface area effects in males at T1 and T2. Associations between tPRS-M and cortical surface area in males are depicted at T1 **(a)** and T2 **(b)**. 10 genetic principle components, eTIV, and study site were regressed out.

**Supplementary Figure 4.**
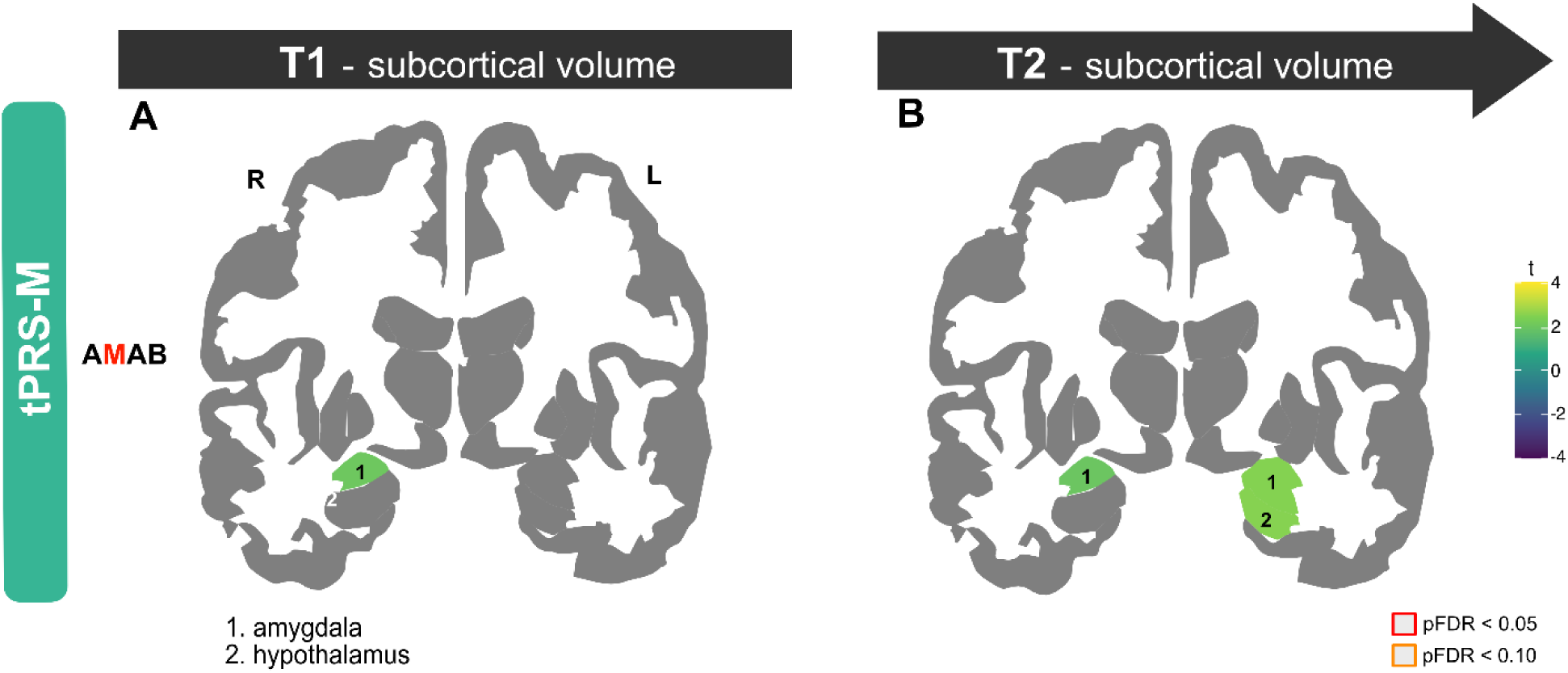
tPRS-M subcortical volume effects in males at T1 and T2. Associations between tPRS-M and subcortical volume in males are depicted at T1 **(a)** and T2 **(b)**. 10 genetic principle components, eTIV, and study site were regressed out.

## Supplementary Tables

**Table S1.** Effects of tPRS-F on depressive symptoms at T1.

| CBCL Syndrome Scale | Main effect, females<br>(n = 2338) |  |  | Main effect, males<br>(n = 2664) |  |  |
| --- | --- | --- | --- | --- | --- | --- |
|  | t | p | pFDR | t | p | pFDR |
| Anxious/Depressed | -0.889 | 0.374 | 0.374 | 0.061 | 0.951 | 0.951 |
| Withdrawn/Depressed | 1.901 | 0.057 | 0.114 | -0.58 | 0.562 | 0.951 |

*FDR = false discovery rate; bold indicates pFDR < 0.05.*

**Table S2.** Effects of tPRS-M on depressive symptoms at T1.

| CBCL Syndrome Scale | Main effect, females<br>(n = 2338) |  |  | Main effect, males<br>(n = 2664) |  |  |
| --- | --- | --- | --- | --- | --- | --- |
|  | t | p | pFDR | t | p | pFDR |
| Anxious/Depressed | -0.928 | 0.354 | 0.708 | **3.192** | **0.001** | **0.002** |
| Withdrawn/Depressed | -0.256 | 0.798 | 0.798 | **2.566** | **0.01** | **0.01** |

*FDR = false discovery rate; bold indicates pFDR < 0.05.*

**Table S3.** Effects of tPRS-F on regional cortical morphology at T1.

| Desikan-Killiany<br>cortical region | Cortical thickness |  |  |  |  |  | Cortical surface area |  |  |  |  |  |
| --- | --- | --- | --- | --- | --- | --- | --- | --- | --- | --- | --- | --- |
|  | Main effect, females |  |  | Main effect, males |  |  | Main effect, females |  |  | Main effect, males |  |  |
|  | (n = 2320) |  |  | (n = 2657) |  |  | (n = 2320) |  |  | (n = 2657) |  |  |
|  | t | p | pFDR | t | p | pFDR | t | p | pFDR | t | p | pFDR |
| Banks superior temporal sulcus, lh | 0.203 | 0.839 | 0.931 | -0.085 | 0.932 | 0.95 | 0.172 | 0.864 | 0.971 | 0.591 | 0.555 | 0.961 |
| Caudal anterior cingulate, lh | 0.084 | 0.933 | 0.961 | 0.834 | 0.404 | 0.939 | 0.171 | 0.864 | 0.971 | 1.3 | 0.194 | 0.961 |
| Caudal middle frontal, lh | 0.118 | 0.906 | 0.961 | -2.111 | 0.035 | 0.854 | 0.447 | 0.655 | 0.971 | 1.251 | 0.211 | 0.961 |
| Cuneus, lh | 0.542 | 0.588 | 0.931 | -0.627 | 0.531 | 0.939 | 0.579 | 0.563 | 0.971 | 0.105 | 0.916 | 0.961 |
| Entorhinal, lh | -0.321 | 0.749 | 0.931 | -0.814 | 0.416 | 0.939 | 1.129 | 0.259 | 0.971 | 0.117 | 0.907 | 0.961 |
| Fusiform, lh | -1.224 | 0.221 | 0.929 | -0.357 | 0.721 | 0.939 | -0.157 | 0.876 | 0.971 | -0.21 | 0.834 | 0.961 |
| Inferior parietal, lh | -0.859 | 0.39 | 0.931 | -0.3 | 0.764 | 0.939 | 1.272 | 0.204 | 0.971 | 0.565 | 0.572 | 0.961 |
| Inferior temporal, lh | -1.16 | 0.246 | 0.929 | 0.099 | 0.921 | 0.95 | 0.822 | 0.411 | 0.971 | -1.826 | 0.068 | 0.688 |
| Isthmus cingulate, lh | -0.483 | 0.629 | 0.931 | 0.773 | 0.439 | 0.939 | -0.424 | 0.672 | 0.971 | 0.915 | 0.36 | 0.961 |
| Lateral occipital, lh | -0.224 | 0.823 | 0.931 | -0.676 | 0.499 | 0.939 | 0.298 | 0.766 | 0.971 | 0.166 | 0.868 | 0.961 |
| Lateral orbitofrontal, lh | 0.255 | 0.799 | 0.931 | 0.361 | 0.718 | 0.939 | 0.418 | 0.676 | 0.971 | -0.102 | 0.919 | 0.961 |
| Lingual, lh | -0.488 | 0.626 | 0.931 | 0.325 | 0.745 | 0.939 | 0.327 | 0.743 | 0.971 | 0.496 | 0.62 | 0.961 |
| Medial orbitofrontal, lh | 0.2 | 0.841 | 0.931 | 0.309 | 0.757 | 0.939 | -1.001 | 0.317 | 0.971 | -1.112 | 0.266 | 0.961 |
| Middle temporal, lh | -0.172 | 0.863 | 0.931 | -0.537 | 0.591 | 0.939 | -0.5 | 0.617 | 0.971 | -1.316 | 0.188 | 0.961 |
| Parahippocampal, lh | 1.161 | 0.246 | 0.929 | -0.306 | 0.759 | 0.939 | -0.328 | 0.743 | 0.971 | 0.89 | 0.374 | 0.961 |
| Paracentral, lh | -0.474 | 0.636 | 0.931 | -1.6 | 0.11 | 0.854 | -0.19 | 0.849 | 0.971 | 0.819 | 0.413 | 0.961 |
| Pars opercularis, lh | -0.628 | 0.53 | 0.931 | 1.261 | 0.207 | 0.939 | -1.996 | 0.046 | 0.971 | -2.613 | 0.009 | 0.578 |
| Pars orbitalis, lh | -1.241 | 0.215 | 0.929 | -0.369 | 0.712 | 0.939 | -0.63 | 0.529 | 0.971 | -0.366 | 0.715 | 0.961 |
| Pars triangularis, lh | 0.598 | 0.55 | 0.931 | 0.264 | 0.792 | 0.939 | -0.921 | 0.357 | 0.971 | -0.51 | 0.61 | 0.961 |
| Pericalcarine, lh | -1.38 | 0.168 | 0.929 | -1.004 | 0.315 | 0.939 | 0.677 | 0.498 | 0.971 | 0.97 | 0.332 | 0.961 |
| Postcentral, lh | -0.22 | 0.826 | 0.931 | -0.546 | 0.585 | 0.939 | -0.141 | 0.888 | 0.971 | -0.532 | 0.595 | 0.961 |
| Posterior cingulate, lh | -0.179 | 0.858 | 0.931 | -1.088 | 0.277 | 0.939 | -1.135 | 0.256 | 0.971 | 0.627 | 0.531 | 0.961 |
| Precentral, lh | -0.86 | 0.39 | 0.931 | -0.663 | 0.507 | 0.939 | -0.108 | 0.914 | 0.971 | 1.185 | 0.236 | 0.961 |
| Precuneus, lh | 0.639 | 0.523 | 0.931 | -0.208 | 0.835 | 0.946 | -0.785 | 0.432 | 0.971 | 0.391 | 0.696 | 0.961 |
| Rostral anterior cingulate,<br>lh | -1.412 | 0.158 | 0.929 | 0.568 | 0.57 | 0.939 | 0.388 | 0.698 | 0.971 | 1.471 | 0.141 | 0.872 |
| Rostral middle frontal, lh | -0.468 | 0.64 | 0.931 | -0.174 | 0.862 | 0.95 | -0.717 | 0.474 | 0.971 | 0.003 | 0.997 | 0.997 |
| Superior frontal, lh | -1.468 | 0.142 | 0.929 | 0.338 | 0.735 | 0.939 | 0.16 | 0.873 | 0.971 | 0.587 | 0.557 | 0.961 |
| Superior parietal, lh | -0.258 | 0.796 | 0.931 | 0.383 | 0.702 | 0.939 | -0.81 | 0.418 | 0.971 | 0.388 | 0.698 | 0.961 |
| Superior temporal, lh | -0.784 | 0.433 | 0.931 | -0.806 | 0.42 | 0.939 | -1.884 | 0.06 | 0.971 | -0.465 | 0.642 | 0.961 |
| Supramarginal, lh | -0.031 | 0.976 | 0.976 | -0.799 | 0.424 | 0.939 | -1.48 | 0.139 | 0.971 | 0.605 | 0.545 | 0.961 |
| Frontal pole, lh | 1.019 | 0.308 | 0.931 | -0.401 | 0.689 | 0.939 | -0.632 | 0.528 | 0.971 | 0.45 | 0.653 | 0.961 |
| Temporal pole, lh | -2.43 | 0.015 | 0.354 | -2.129 | 0.033 | 0.854 | -0.41 | 0.682 | 0.971 | 0.973 | 0.33 | 0.961 |
| Transverse temporal, lh | -1.306 | 0.192 | 0.929 | -0.738 | 0.461 | 0.939 | 0.011 | 0.991 | 0.991 | -0.575 | 0.565 | 0.961 |
| Insula, lh | -0.936 | 0.349 | 0.931 | -1.2 | 0.23 | 0.939 | -0.99 | 0.323 | 0.971 | 1.688 | 0.091 | 0.688 |
| Banks superior temporal sulcus, rh | 0.275 | 0.783 | 0.931 | -0.434 | 0.664 | 0.939 | -0.174 | 0.862 | 0.971 | 0.063 | 0.95 | 0.977 |
| Caudal anterior cingulate, rh | 0.416 | 0.677 | 0.931 | -1.817 | 0.069 | 0.854 | 1.429 | 0.153 | 0.971 | -1.806 | 0.071 | 0.688 |
| Caudal middle frontal, rh | -2.368 | 0.018 | 0.354 | -0.238 | 0.812 | 0.939 | -0.278 | 0.781 | 0.971 | -0.813 | 0.416 | 0.961 |
| Cuneus, rh | 0.277 | 0.782 | 0.931 | -0.524 | 0.6 | 0.939 | -0.346 | 0.729 | 0.971 | -1.09 | 0.276 | 0.961 |
| Entorhinal, rh | -0.69 | 0.49 | 0.931 | 1.61 | 0.107 | 0.854 | 0.072 | 0.943 | 0.972 | -1.857 | 0.063 | 0.688 |
| Fusiform, rh | -1.272 | 0.204 | 0.929 | -0.019 | 0.985 | 0.985 | -0.174 | 0.862 | 0.971 | 0.309 | 0.757 | 0.961 |
| Inferior parietal, rh | 0.609 | 0.543 | 0.931 | -0.11 | 0.913 | 0.95 | -0.902 | 0.367 | 0.971 | 0.205 | 0.838 | 0.961 |
| Inferior temporal, rh | -1.083 | 0.279 | 0.931 | -0.665 | 0.506 | 0.939 | 0.6 | 0.548 | 0.971 | -0.972 | 0.331 | 0.961 |
| Isthmus cingulate, rh | 0.356 | 0.722 | 0.931 | 0.591 | 0.555 | 0.939 | -0.622 | 0.534 | 0.971 | -0.455 | 0.649 | 0.961 |
| Lateral occipital, rh | -0.087 | 0.931 | 0.961 | -0.932 | 0.352 | 0.939 | -0.313 | 0.754 | 0.971 | 0.415 | 0.678 | 0.961 |
| Lateral orbitofrontal, rh | -0.32 | 0.749 | 0.931 | -0.768 | 0.443 | 0.939 | -0.141 | 0.888 | 0.971 | -0.154 | 0.878 | 0.961 |
| Lingual, rh | -0.696 | 0.486 | 0.931 | -0.404 | 0.686 | 0.939 | -0.182 | 0.856 | 0.971 | 0.047 | 0.963 | 0.977 |
| Medial orbitofrontal, rh | 0.639 | 0.523 | 0.931 | 1.195 | 0.232 | 0.939 | -0.282 | 0.778 | 0.971 | 0.398 | 0.691 | 0.961 |
| Middle temporal, rh | -0.847 | 0.397 | 0.931 | -1.674 | 0.094 | 0.854 | -0.608 | 0.543 | 0.971 | -0.124 | 0.901 | 0.961 |
| Parahippocampal, rh | 0.055 | 0.956 | 0.97 | -1.586 | 0.113 | 0.854 | 0.354 | 0.723 | 0.971 | 0.521 | 0.602 | 0.961 |
| Paracentral, rh | -0.815 | 0.415 | 0.931 | -1.037 | 0.3 | 0.939 | 1.509 | 0.132 | 0.971 | 0.896 | 0.37 | 0.961 |
| Pars opercularis, rh | -2.234 | 0.026 | 0.354 | -0.116 | 0.907 | 0.95 | 0.667 | 0.505 | 0.971 | -1.563 | 0.118 | 0.802 |
| Pars orbitalis, rh | -0.826 | 0.409 | 0.931 | -0.08 | 0.936 | 0.95 | -1.283 | 0.2 | 0.971 | 0.786 | 0.432 | 0.961 |
| Pars triangularis, rh | -1.127 | 0.26 | 0.931 | -0.311 | 0.756 | 0.939 | -0.571 | 0.568 | 0.971 | 0.602 | 0.547 | 0.961 |
| Pericalcarine, rh | -0.726 | 0.468 | 0.931 | 0.52 | 0.603 | 0.939 | 0.628 | 0.53 | 0.971 | 0.198 | 0.843 | 0.961 |
| Postcentral, rh | -0.514 | 0.607 | 0.931 | -0.118 | 0.906 | 0.95 | 0.09 | 0.928 | 0.971 | 0.403 | 0.687 | 0.961 |
| Posterior cingulate, rh | 0.425 | 0.671 | 0.931 | 0.234 | 0.815 | 0.939 | -0.124 | 0.901 | 0.971 | -0.802 | 0.423 | 0.961 |
| Precentral, rh | -2.25 | 0.025 | 0.354 | -1.63 | 0.103 | 0.854 | -1.52 | 0.129 | 0.971 | 0.167 | 0.867 | 0.961 |
| Precuneus, rh | -0.23 | 0.818 | 0.931 | -0.652 | 0.514 | 0.939 | -1.514 | 0.13 | 0.971 | -0.142 | 0.887 | 0.961 |
| Rostral anterior cingulate, rh | 0.331 | 0.741 | 0.931 | 0.357 | 0.721 | 0.939 | -0.24 | 0.811 | 0.971 | -1.744 | 0.081 | 0.688 |
| Rostral middle frontal, rh | -1.67 | 0.095 | 0.807 | 1.183 | 0.237 | 0.939 | 0.858 | 0.391 | 0.971 | -0.971 | 0.332 | 0.961 |
| Superior frontal, rh | -1.821 | 0.069 | 0.67 | 0.439 | 0.661 | 0.939 | 0.796 | 0.426 | 0.971 | 1.723 | 0.085 | 0.688 |
| Superior parietal, rh | -0.385 | 0.7 | 0.931 | -0.243 | 0.808 | 0.939 | -0.929 | 0.353 | 0.971 | -0.157 | 0.876 | 0.961 |
| Superior temporal, rh | -1.546 | 0.122 | 0.922 | -1.29 | 0.197 | 0.939 | -0.052 | 0.959 | 0.973 | -0.178 | 0.859 | 0.961 |
| Supramarginal, rh | -0.982 | 0.326 | 0.931 | -0.616 | 0.538 | 0.939 | 0.141 | 0.888 | 0.971 | 2.391 | 0.017 | 0.578 |
| Frontal pole, rh | -1.034 | 0.301 | 0.931 | -0.853 | 0.394 | 0.939 | -0.808 | 0.419 | 0.971 | 1.922 | 0.055 | 0.688 |
| Temporal pole, rh | -2.738 | 0.006 | 0.354 | -2.016 | 0.044 | 0.854 | -0.19 | 0.849 | 0.971 | -0.278 | 0.781 | 0.961 |
| Transverse temporal, rh | -0.329 | 0.742 | 0.931 | -0.999 | 0.318 | 0.939 | 1.807 | 0.071 | 0.971 | -0.109 | 0.913 | 0.961 |
| Insula, rh | -2.105 | 0.035 | 0.397 | -0.493 | 0.622 | 0.939 | -0.193 | 0.847 | 0.971 | 0.919 | 0.358 | 0.961 |

*FDR = false discovery rate; lh = left hemisphere; rh = right hemisphere; bold indicates $pFDR < 0.05$ .*

**Table S4.** Effects of tPRS-F on regional subcortical volume at T1.

| Region | Main effect, females<br>(n = 2320) |  |  | Main effect, males<br>(n = 2657) |  |  |
| --- | --- | --- | --- | --- | --- | --- |
|  | t | p | pFDR | t | p | pFDR |
| Thalamus, lh | -1.931 | 0.054 | 0.126 | 0.632 | 0.527 | 0.665 |
| Caudate, lh | -0.905 | 0.365 | 0.426 | 1.564 | 0.118 | 0.395 |
| Putamen, lh | -0.765 | 0.444 | 0.478 | -0.568 | 0.57 | 0.665 |
| Pallidum, lh | -1.755 | 0.079 | 0.158 | 0.615 | 0.539 | 0.665 |
| Hippocampus, lh | -2.568 | 0.01 | 0.126 | 1.097 | 0.273 | 0.548 |
| Amygdala, lh | -2.091 | 0.037 | 0.126 | -0.446 | 0.656 | 0.706 |
| Accumbens area,<br>lh | -1.625 | 0.104 | 0.182 | 1.73 | 0.084 | 0.395 |
| Thalamus, rh | -2.085 | 0.037 | 0.126 | 0.185 | 0.853 | 0.853 |
| Caudate, rh | -2.005 | 0.045 | 0.126 | 1.473 | 0.141 | 0.395 |
| Putamen, rh | -0.427 | 0.67 | 0.67 | -1.073 | 0.283 | 0.548 |
| Pallidum, rh | -1.226 | 0.221 | 0.281 | 1.685 | 0.092 | 0.395 |
| Hippocampus, rh | -2.174 | 0.03 | 0.126 | 0.768 | 0.442 | 0.665 |
| Amygdala, rh | -1.453 | 0.146 | 0.227 | -1.914 | 0.056 | 0.395 |
| Accumbens area,<br>rh | -1.382 | 0.167 | 0.234 | 1.01 | 0.313 | 0.548 |

*FDR = false discovery rate; lh = left hemisphere; rh = right hemisphere; bold indicates pFDR < 0.05.*

**Table S5.** tPRS-F-by-time interaction effects on SCV in females.

| Region | tPRS-F-by-time<br>interaction effect, in<br>females<br>(n = 1584) |  |
| --- | --- | --- |
|  | t | p |
| **Hippocampus, lh** | **-2.247** | **0.025** |
| Caudate, rh | 0.683 | 0.495 |
| Hippocampus, rh | -1.413 | 0.158 |
| Accumbens area, rh | -1.648 | 0.099 |

*lh = left hemisphere; rh = right hemisphere; bold indicates $p < 0.05$ .*

**Table S6.** Effects of tPRS-M on regional cortical morphology at T1.

| Desikan-Killiany<br>cortical region | Cortical thickness |  |  |  |  |  | Cortical surface area |  |  |  |  |  |
| --- | --- | --- | --- | --- | --- | --- | --- | --- | --- | --- | --- | --- |
|  | Main effect, females |  |  | Main effect, males |  |  | Main effect, females |  |  | Main effect, males |  |  |
|  | (n = 2320) |  |  | (n = 2657) |  |  | (n = 2320) |  |  | (n = 2657) |  |  |
|  | t | p | pFDR | t | p | pFDR | t | p | pFDR | t | p | pFDR |
| Banks superior temporal sulcus, lh | -0.882 | 0.378 | 0.941 | -1.651 | 0.099 | 0.277 | -1.879 | 0.06 | 0.544 | -2.417 | 0.016 | 0.612 |
| Caudal anterior cingulate, lh | 0.23 | 0.818 | 0.979 | 0.781 | 0.435 | 0.689 | -0.499 | 0.618 | 0.934 | 0.004 | 0.997 | 0.997 |
| Caudal middle frontal, lh | 0.75 | 0.453 | 0.941 | 2.358 | 0.018 | 0.408 | -1.107 | 0.269 | 0.899 | 1.171 | 0.242 | 0.904 |
| Cuneus, lh | 0.009 | 0.993 | 0.993 | 1.259 | 0.208 | 0.615 | -1.856 | 0.064 | 0.544 | 0.004 | 0.997 | 0.997 |
| Entorhinal, lh | 1.3 | 0.194 | 0.941 | -0.136 | 0.892 | 0.905 | -0.603 | 0.547 | 0.934 | -0.583 | 0.56 | 0.904 |
| Fusiform, lh | 0.628 | 0.53 | 0.941 | 0.846 | 0.398 | 0.682 | 0.084 | 0.933 | 0.975 | -1.097 | 0.273 | 0.904 |
| Inferior parietal, lh | -0.536 | 0.592 | 0.941 | 1.299 | 0.194 | 0.615 | -0.261 | 0.794 | 0.971 | -0.479 | 0.632 | 0.904 |
| Inferior temporal, lh | 0.222 | 0.824 | 0.979 | 0.333 | 0.739 | 0.843 | -0.586 | 0.558 | 0.934 | 0.141 | 0.888 | 0.957 |
| Isthmus cingulate, lh | -0.029 | 0.977 | 0.993 | 0.977 | 0.329 | 0.676 | -0.303 | 0.762 | 0.971 | -0.138 | 0.89 | 0.957 |
| Lateral occipital, lh | 0.665 | 0.506 | 0.941 | 1.579 | 0.114 | 0.615 | -1.558 | 0.119 | 0.649 | 0.452 | 0.651 | 0.904 |
| Lateral orbitofrontal, lh | -1.638 | 0.102 | 0.941 | 1.03 | 0.303 | 0.676 | -0.032 | 0.975 | 0.975 | -0.498 | 0.618 | 0.904 |
| Lingual, lh | 1.127 | 0.26 | 0.941 | 1.144 | 0.253 | 0.676 | -1.223 | 0.222 | 0.899 | 0.671 | 0.502 | 0.904 |
| Medial orbitofrontal, lh | -0.992 | 0.321 | 0.941 | -0.642 | 0.521 | 0.738 | 0.436 | 0.663 | 0.959 | 0.632 | 0.527 | 0.904 |
| Middle temporal, lh | 0.601 | 0.548 | 0.941 | 0.839 | 0.401 | 0.682 | -1.884 | 0.06 | 0.544 | -1.338 | 0.181 | 0.904 |
| Parahippocampal, lh | 1.366 | 0.172 | 0.941 | -0.268 | 0.789 | 0.865 | 1.539 | 0.124 | 0.649 | -1.292 | 0.196 | 0.904 |
| Paracentral, lh | 0.753 | 0.451 | 0.941 | 0.362 | 0.718 | 0.843 | -0.263 | 0.793 | 0.971 | -0.065 | 0.948 | 0.992 |
| Pars opercularis, lh | -1.299 | 0.194 | 0.941 | 1.437 | 0.151 | 0.615 | 0.503 | 0.615 | 0.934 | 0.552 | 0.581 | 0.904 |
| Pars orbitalis, lh | -1.087 | 0.277 | 0.941 | 0.996 | 0.319 | 0.676 | -2.074 | 0.038 | 0.544 | -0.246 | 0.806 | 0.945 |
| Pars triangularis, lh | -0.085 | 0.932 | 0.993 | 0.387 | 0.699 | 0.843 | 0.56 | 0.576 | 0.934 | 0.603 | 0.546 | 0.904 |
| Pericalcarine, lh | 0.905 | 0.366 | 0.941 | 0.515 | 0.606 | 0.792 | -1.628 | 0.104 | 0.643 | 1.114 | 0.265 | 0.904 |
| Postcentral, lh | 0.22 | 0.826 | 0.979 | 1.447 | 0.148 | 0.615 | -0.611 | 0.542 | 0.934 | 0.809 | 0.419 | 0.904 |
| Posterior cingulate, lh | 1.406 | 0.16 | 0.941 | 0.306 | 0.76 | 0.847 | 0.147 | 0.883 | 0.971 | 0.125 | 0.901 | 0.957 |
| Precentral, lh | 0.046 | 0.963 | 0.993 | 2.813 | 0.005 | 0.34 | -1.081 | 0.28 | 0.899 | -0.53 | 0.596 | 0.904 |
| Precuneus, lh | 0.995 | 0.32 | 0.941 | 0.443 | 0.658 | 0.816 | -0.516 | 0.606 | 0.934 | 1.317 | 0.188 | 0.904 |
| Rostral anterior cingulate, lh | 0.225 | 0.822 | 0.979 | 1.431 | 0.153 | 0.615 | 0.646 | 0.518 | 0.934 | 0.339 | 0.734 | 0.925 |
| Rostral middle frontal, lh | 1.699 | 0.09 | 0.941 | 0.519 | 0.604 | 0.792 | -1.135 | 0.257 | 0.899 | 0.714 | 0.475 | 0.904 |
| Superior frontal, lh | 0.071 | 0.943 | 0.993 | 0.202 | 0.84 | 0.886 | -1.631 | 0.103 | 0.643 | -1.078 | 0.281 | 0.904 |
| Superior parietal, lh | -0.191 | 0.849 | 0.979 | 0.439 | 0.66 | 0.816 | 0.189 | 0.85 | 0.971 | -1.03 | 0.303 | 0.904 |
| Superior temporal, lh | 0.549 | 0.583 | 0.941 | -0.052 | 0.958 | 0.958 | -1.102 | 0.271 | 0.899 | -1.474 | 0.141 | 0.904 |
| Supramarginal, lh | 1.031 | 0.303 | 0.941 | 0.335 | 0.738 | 0.843 | -0.185 | 0.854 | 0.971 | -1.593 | 0.111 | 0.904 |
| Frontal pole, lh | -0.356 | 0.722 | 0.979 | 0.748 | 0.454 | 0.689 | -0.23 | 0.818 | 0.971 | -0.15 | 0.881 | 0.957 |
| Temporal pole, lh | 0.876 | 0.381 | 0.941 | 0.749 | 0.454 | 0.689 | 0.681 | 0.496 | 0.934 | -1.192 | 0.234 | 0.904 |
| Transverse temporal, lh | 0.197 | 0.844 | 0.979 | 1.307 | 0.191 | 0.615 | -0.967 | 0.333 | 0.899 | 0.678 | 0.498 | 0.904 |
| Insula, lh | 0.702 | 0.483 | 0.941 | 0.919 | 0.358 | 0.676 | -0.152 | 0.879 | 0.971 | 1.495 | 0.135 | 0.904 |
| Banks superior temporal sulcus, rh | 0.417 | 0.677 | 0.979 | 0.608 | 0.543 | 0.754 | -0.755 | 0.451 | 0.934 | -0.453 | 0.651 | 0.904 |
| Caudal anterior cingulate, rh | -0.532 | 0.595 | 0.941 | 1.099 | 0.272 | 0.676 | -1.108 | 0.268 | 0.899 | -1.245 | 0.213 | 0.904 |
| Caudal middle frontal, rh | 0.711 | 0.477 | 0.941 | 1.269 | 0.204 | 0.615 | 0.502 | 0.616 | 0.934 | 1.394 | 0.163 | 0.904 |
| Cuneus, rh | 1.086 | 0.278 | 0.941 | 0.867 | 0.386 | 0.682 | -1.985 | 0.047 | 0.544 | 0.007 | 0.995 | 0.997 |
| Entorhinal, rh | 0.194 | 0.846 | 0.979 | 0.141 | 0.888 | 0.905 | 0.313 | 0.754 | 0.971 | -0.398 | 0.69 | 0.92 |
| Fusiform, rh | 1.082 | 0.279 | 0.941 | 0.708 | 0.479 | 0.693 | 0.195 | 0.846 | 0.971 | -0.659 | 0.51 | 0.904 |
| Inferior parietal, rh | -0.59 | 0.555 | 0.941 | 1 | 0.317 | 0.676 | -0.227 | 0.82 | 0.971 | -0.317 | 0.751 | 0.925 |
| Inferior temporal, rh | 0.624 | 0.533 | 0.941 | -0.247 | 0.805 | 0.869 | -0.857 | 0.391 | 0.907 | 0.257 | 0.797 | 0.945 |
| Isthmus cingulate, rh | 0.606 | 0.545 | 0.941 | 1.489 | 0.137 | 0.615 | 0.123 | 0.902 | 0.971 | -0.184 | 0.854 | 0.957 |
| Lateral occipital, rh | 0.785 | 0.433 | 0.941 | 2.139 | 0.033 | 0.449 | -0.889 | 0.374 | 0.907 | 0.831 | 0.406 | 0.904 |
| Lateral orbitofrontal, rh | -0.671 | 0.503 | 0.941 | 1.431 | 0.153 | 0.615 | 0.129 | 0.897 | 0.971 | -0.434 | 0.665 | 0.904 |
| Lingual, rh | 0.682 | 0.495 | 0.941 | 1.868 | 0.062 | 0.615 | -0.952 | 0.341 | 0.899 | 0.786 | 0.432 | 0.904 |
| Medial orbitofrontal, rh | -1.324 | 0.186 | 0.941 | -0.462 | 0.644 | 0.816 | 0.416 | 0.677 | 0.959 | -0.317 | 0.751 | 0.925 |
| Middle temporal, rh | 0.685 | 0.493 | 0.941 | 1.771 | 0.077 | 0.615 | -0.71 | 0.478 | 0.934 | -1.606 | 0.108 | 0.904 |
| Parahippocampal, rh | 0.419 | 0.675 | 0.979 | -0.327 | 0.744 | 0.843 | -1.043 | 0.297 | 0.899 | -0.675 | 0.5 | 0.904 |
| Paracentral, rh | -0.194 | 0.846 | 0.979 | 0.932 | 0.351 | 0.676 | -0.842 | 0.4 | 0.907 | -0.739 | 0.46 | 0.904 |
| Pars opercularis, rh | 0.231 | 0.817 | 0.979 | 2.368 | 0.018 | 0.408 | 1.145 | 0.252 | 0.899 | -0.525 | 0.6 | 0.904 |
| Pars orbitalis, rh | 0.253 | 0.801 | 0.979 | -0.756 | 0.45 | 0.689 | -0.94 | 0.348 | 0.899 | -0.696 | 0.486 | 0.904 |
| Pars triangularis, rh | 0.653 | 0.514 | 0.941 | 1.594 | 0.111 | 0.615 | -0.921 | 0.357 | 0.899 | -0.869 | 0.385 | 0.904 |
| Pericalcarine, rh | 1.053 | 0.293 | 0.941 | 0.529 | 0.597 | 0.792 | -1.649 | 0.099 | 0.643 | 0.511 | 0.61 | 0.904 |
| Postcentral, rh | -0.547 | 0.584 | 0.941 | 1.103 | 0.27 | 0.676 | 0.052 | 0.959 | 0.975 | -0.135 | 0.893 | 0.957 |
| Posterior cingulate, rh | -0.836 | 0.403 | 0.941 | 0.874 | 0.382 | 0.682 | -0.506 | 0.613 | 0.934 | -1.532 | 0.126 | 0.904 |
| Precentral, rh | 0.827 | 0.408 | 0.941 | 2.243 | 0.025 | 0.425 | -0.231 | 0.818 | 0.971 | 0.358 | 0.72 | 0.925 |
| Precuneus, rh | 0.069 | 0.945 | 0.993 | 0.193 | 0.847 | 0.886 | 0.035 | 0.972 | 0.975 | 1.066 | 0.286 | 0.904 |
| Rostral anterior cingulate, rh | -0.073 | 0.942 | 0.993 | 1.048 | 0.295 | 0.676 | 0.63 | 0.529 | 0.934 | -0.303 | 0.762 | 0.925 |
| Rostral middle frontal, rh | 0.509 | 0.611 | 0.944 | 1.231 | 0.218 | 0.618 | -2.199 | 0.028 | 0.544 | 0.437 | 0.662 | 0.904 |
| Superior frontal, rh | 0.806 | 0.421 | 0.941 | 0.711 | 0.477 | 0.693 | -1.492 | 0.136 | 0.661 | 0.594 | 0.553 | 0.904 |
| Superior parietal, rh | 0.442 | 0.659 | 0.979 | 1.319 | 0.187 | 0.615 | 0.974 | 0.33 | 0.899 | -0.951 | 0.342 | 0.904 |
| Superior temporal, rh | 0.258 | 0.796 | 0.979 | 1.426 | 0.154 | 0.615 | -0.45 | 0.653 | 0.959 | -2.376 | 0.018 | 0.612 |
| Supramarginal, rh | 0.009 | 0.993 | 0.993 | 1.033 | 0.302 | 0.676 | 2.025 | 0.043 | 0.544 | -1.59 | 0.112 | 0.904 |
| Frontal pole, rh | -0.021 | 0.983 | 0.993 | -0.746 | 0.456 | 0.689 | -2.4 | 0.016 | 0.544 | 0.48 | 0.631 | 0.904 |
| Temporal pole, rh | 0.808 | 0.419 | 0.941 | 0.941 | 0.347 | 0.676 | 0.191 | 0.849 | 0.971 | 0.951 | 0.342 | 0.904 |
| Transverse temporal, rh | -0.949 | 0.343 | 0.941 | 1.72 | 0.086 | 0.615 | -0.109 | 0.914 | 0.971 | -1.009 | 0.313 | 0.904 |
| Insula, rh | -0.662 | 0.508 | 0.941 | 1.374 | 0.17 | 0.615 | -0.699 | 0.485 | 0.934 | -0.528 | 0.598 | 0.904 |

*FDR = false discovery rate; lh = left hemisphere; rh = right hemisphere; bold indicates $pFDR < 0.05$ .*

**Table S7.** Effects of tPRS-M on regional subcortical volume at T1.

| Region | Main effect, females<br>(n = 2320) |  |  | Main effect, males<br>(n = 2657) |  |  |
| --- | --- | --- | --- | --- | --- | --- |
|  | t | p | pFDR | t | p | pFDR |
| Thalamus, lh | -1.21 | 0.226 | 0.638 | 0.005 | 0.996 | 0.996 |
| Caudate, lh | 0.574 | 0.566 | 0.638 | -1.199 | 0.231 | 0.404 |
| Putamen, lh | 1.489 | 0.137 | 0.638 | -0.68 | 0.497 | 0.696 |
| Pallidum, lh | 0.467 | 0.64 | 0.64 | 0.928 | 0.353 | 0.549 |
| Hippocampus, lh | -0.772 | 0.44 | 0.638 | 1.702 | 0.089 | 0.277 |
| Amygdala, lh | 1.312 | 0.19 | 0.638 | 2.042 | 0.041 | 0.277 |
| Accumbens area,<br>lh | 0.858 | 0.391 | 0.638 | -1.561 | 0.119 | 0.278 |
| Thalamus, rh | -0.721 | 0.471 | 0.638 | 0.298 | 0.766 | 0.956 |
| Caudate, rh | 0.761 | 0.447 | 0.638 | -1.708 | 0.088 | 0.277 |
| Putamen, rh | 1.385 | 0.166 | 0.638 | -0.14 | 0.888 | 0.956 |
| Pallidum, rh | 0.548 | 0.584 | 0.638 | -0.218 | 0.827 | 0.956 |
| Hippocampus, rh | -0.537 | 0.592 | 0.638 | 1.703 | 0.089 | 0.277 |
| Amygdala, rh | 0.582 | 0.561 | 0.638 | 1.273 | 0.203 | 0.404 |
| Accumbens area,<br>rh | 0.992 | 0.321 | 0.638 | 0.005 | 0.996 | 0.996 |

*FDR = false discovery rate; lh = left hemisphere; rh = right hemisphere; bold indicates pFDR < 0.05*

## References

1. American Psychiatric Association. Diagnostic and statistical manual of mental disorders. 5th ed. Washington, D.C.; 2013.

2. Kalin NH. The critical relationship between anxiety and depression. AJP. 2020;177:365–367.

3. Brown TA, Campbell LA, Lehman CL, Grisham JR, Mancill RB. Current and lifetime comorbidity of the DSM-IV anxiety and mood disorders in a large clinical sample. Journal of Abnormal Psychology. 2001;110:585–599.

4. Judd LL, Kessler RC, PauIus MP, Zeller PV, Wittchen H-U., Kunovac JL. Comorbidity as a fundamental feature of generalized anxiety disorders: Results from the National Comorbidity Study (NCS). Acta Psychiatr Scand. 1998;98:6–11.

5. Kessler RC, Berglund P, Demler O, Jin R, Merikangas KR, Walters EE. Lifetime prevalence and age-of-onset distributions of DSM-IV disorders in the national comorbidity survey replication. Arch Gen Psychiatry. 2005 Jun;62(6):593–602.

6. Kornstein SG, Schatzberg AF, Thase ME, Yonkers KA, McCullough JP, Keitner GI, et al. Gender differences in chronic major and double depression. J Affect Disord. 2000 Oct;60(1):1–11.

7. Angst J, Dobler-Mikola A. Do the diagnostic criteria determine the sex ratio in depression? J Affect Disord. 1984 Dec 1;7(3):189–98.

8. Young MA, Fogg LF, Scheftner WA, Keller MB, Fawcett JA. Sex differences in the lifetime prevalence of depression: does varying the diagnostic criteria reduce the female/male ratio? J Affect Disord. 1990 Mar 1;18(3):187–92.

9. Scheibe S, Preuschhof C, Cristi C, Bagby RM. Are there gender differences in major depression and its response to antidepressants? J Affect Disord. 2003 Aug;75(3):223– 35.

10. Burcusa SL, Iacono WG. Risk for recurrence in depression. Clin Psychol Rev. 2007 Dec;27(8):959–85.

11. Kessler RC. Epidemiology of women and depression. J Affect Disord. 2003 Mar 1;74(1):5–13.

12. Goodwin RD, Gotlib IH. Gender differences in depression: the role of personality factors. Psychiatry Res. 2004 Apr 30;126(2):135–42.

13. Bulloch AGM, Williams JVA, Lavorato DH, Patten SB. The depression and marital status relationship is modified by both age and gender. J Affect Disord. 2017 Dec 1;223:65–8.

14. Kuehner C. Why is depression more common among women than among men? Lancet Psychiatry. 2017 Feb 1;4(2):146–58.

15. Seney ML, Huo Z, Cahill K, French L, Puralewski R, Zhang J, et al. Opposite molecular signatures of depression in men and women. Biol Psychiatry. 2018 Jul 1;84(1):18–27.

16. Labonté B, Engmann O, Purushothaman I, Menard C, Wang J, Tan C, et al. Sex-specific transcriptional signatures in human depression. Nat Med. 2017 Sep;23(9):1102–11.

17. Mareckova K, Hawco C, Dos Santos FC, Bakht A, Calarco N, Miles AE, et al. Novel polygenic risk score as a translational tool linking depression-related changes in the corticolimbic transcriptome with neural face processing and anhedonic symptoms. Transl Psychiatry. 2020 Nov 24;10:410.

18. Miles AE, Dos Santos FC, Byrne EM, Renteria ME, McIntosh AM, Adams MJ, et al. Transcriptome-based polygenic score links depression-related corticolimbic gene expression changes to sex-specific brain morphology and depression risk. Neuropsychopharmacol Off Publ Am Coll Neuropsychopharmacol. 2021 Dec;46(13):2304–11.

19. Miles AE, Rashid SS, Dos Santos FC, Clifford KP, Sibille E, Nikolova YS. Neurodevelopmental signature of a transcriptome-based polygenic risk score for depression. Psychiatry Res. 2024 Jun; 339:116030.

20. Wray NR, Ripke S, Mattheisen M, Trzaskowski M, Byrne EM, Abdellaoui A, et al. Genome-wide association analyses identify 44 risk variants and refine the genetic architecture of major depression. Nat Genet. 2018 May;50(5):668–81.

21. Nolen-Hoeksema S, Girgus JS. The emergence of gender differences in depression during adolescence. Psychol Bull. 1994 May;115(3):424–43.

22. Paus T, Keshavan M, Giedd JN. Why do many psychiatric disorders emerge during adolescence? Nat Rev Neurosci. 2008 Dec;9(12):947–57.

23. Sisk CL, Foster DL. The neural basis of puberty and adolescence. Nat Neurosci. 2004 Oct;7(10):1040–7.

24. Shi L, Zhang Z, Su B. Sex biased gene expression profiling of human brains at major developmental stages. Sci Rep. 2016 Feb 16;6:21181.

25. Athira KV, Bandopadhyay S, Samudrala PK, Naidu VGM, Lahkar M, Chakravarty S. An Overview of the Heterogeneity of Major Depressive Disorder: Current Knowledge and Future Prospective. CN. 2020;18:168–187.

26. Xu C, Li Q, Efimova O, He L, Tatsumoto S, Stepanova V, et al. Human-specific features of spatial gene expression and regulation in eight brain regions. Genome Res. 2018;28:1097–1110.

27. Cai N, Revez JA, Adams MJ, Andlauer TFM, Breen G, Byrne EM, et al. Minimal phenotyping yields genome-wide association signals of low specificity for major depression. Nat Genet. 2020;52:437–447.

28. Sullivan PF, Neale MC, Kendler KS. Genetic epidemiology of major depression: review and meta-analysis. Am J Psychiatry. 2000 Oct;157(10):1552–62.

29. Kendler KS, Ohlsson H, Lichtenstein P, Sundquist J, Sundquist K. The genetic epidemiology of treated major depression in Sweden. AJP. 2018;175:1137–1144.

30. Mueller TI, Leon AC, Keller MB, Solomon DA, Endicott J, Coryell W, et al. Recurrence after recovery from major depressive disorder during 15 years of observational follow-up. Am J Psychiatry. 1999 Jul;156(7):1000–6.

31. Klein DN, Shankman SA, Lewinsohn PM, Rohde P, Seeley JR. Family study of chronic depression in a community sample of young adults. Am J Psychiatry. 2004 Apr;161(4):646–53.

32. Garavan H, Bartsch H, Conway K, Decastro A, Goldstein RZ, Heeringa S, et al. Recruiting the ABCD sample: Design considerations and procedures. Developmental Cognitive Neuroscience. 2018;32:16–22.

33. Ding Y, Chang LC, Wang X, Guilloux JP, Parrish J, Oh H, et al. Molecular and genetic characterization of depression: overlap with other psychiatric disorders and aging. Complex Psychiatry. 2015;1(1):1–12.

34. Wainberg M, Jacobs GR, Voineskos AN, Tripathy SJ. Neurobiological, familial and genetic risk factors for dimensional psychopathology in the Adolescent Brain Cognitive Development study. Mol Psychiatry. 2022 Jun;27(6):2731–41.

35. Uban KA, Horton MK, Jacobus J, Heyser C, Thompson WK, Tapert SF, et al. Biospecimens and the ABCD study: Rationale, methods of collection, measurement and early data. Dev Cogn Neurosci. 2018 Aug 1;32:97–106.

36. Cunningham F, Allen JE, Allen J, Alvarez-Jarreta J, Amode MR, Armean IM, et al. Ensembl 2022. Nucleic Acids Res. 2022 Jan 7;50(D1):D988–95.

37. Hoffman GE, Bendl J, Voloudakis G, Montgomery KS, Sloofman L, Wang YC, et al. CommonMind Consortium provides transcriptomic and epigenomic data for Schizophrenia and Bipolar Disorder. Sci Data. 2019 Sep 24;6(1):180.

38. Barbeira AN, Dickinson SP, Bonazzola R, Zheng J, Wheeler HE, Torres JM, et al. Exploring the phenotypic consequences of tissue specific gene expression variation inferred from GWAS summary statistics. Nat Commun. 2018 May 8;9(1):1825.

39. Hagler DJ, Hatton SN, Cornejo MD, Makowski C, Fair DA, Dick AS, et al. Image processing and analysis methods for the adolescent brain cognitive development study. NeuroImage. 2019 Nov 15;202:116091.

40. Desikan RS, Ségonne F, Fischl B, Quinn BT, Dickerson BC, Blacker D, et al. An automated labeling system for subdividing the human cerebral cortex on MRI scans into gyral based regions of interest. NeuroImage. 2006 Jul 1;31(3):968–80.

41. Barch DM, Albaugh MD, Avenevoli S, Chang L, Clark DB, Glantz MD, et al. Demographic, physical and mental health assessments in the adolescent brain and cognitive development study: Rationale and description. Dev Cogn Neurosci. 2018 Aug 1;32:55–66.

42. Achenbach TM, Rescorla LA. Manual for the ASEBA school-age forms & profiles: an integrated system of multi-informant assessment. ASEBA; 2001. 238 p.

43. Piovesana A, Senior G. How small is big: sample size and skewness. Assessment. 2018 Sep;25(6):793–800.

44. Campbell S, Marriott M, Nahmias C, MacQueen GM. Lower hippocampal volume in patients suffering from depression: a meta-analysis. Am J Psychiatry. 2004 Apr;161(4):598–607.

45. Rolls ET. The storage and recall of memories in the hippocampo-cortical system. Cell Tissue Res. 2018 Sep;373(3):577–604.

46. Jankord R, Herman JP. Limbic regulation of hypothalamo-pituitary-adrenocortical function during acute and chronic stress. Ann N Y Acad Sci. 2008 Dec;1148:64–73.

47. Schmaal L, Veltman DJ, van Erp TGM, Sämann PG, Frodl T, Jahanshad N, et al. Subcortical brain alterations in major depressive disorder: findings from the ENIGMA Major Depressive Disorder working group. Mol Psychiatry. 2016 Jun;21(6):806–12.

48. Videbech P, Ravnkilde B. Hippocampal volume and depression: a meta-analysis of MRI studies. Am J Psychiatry. 2004 Nov;161(11):1957–66.

49. Naninck EFG, Lucassen PJ, Bakker J. Sex differences in adolescent depression: do sex hormones determine vulnerability? J Neuroendocrinol. 2011 May;23(5):383–92.

50. Herman JP, McKlveen JM, Ghosal S, Kopp B, Wulsin A, Makinson R, et al. Regulation of the hypothalamic-pituitary-adrenocortical stress response. Compr Physiol. 2016;6(2):603.

51. Whittle S, Lichter R, Dennison M, Vijayakumar N, Schwartz O, Byrne ML, et al. Structural brain development and depression onset during adolescence: a prospective longitudinal study. Am J Psychiatry. 2014 May;171(5):564–71.

52. Baumann B, Krell D, Dobrowolny H, Bielau H. Mechanisms of action in the prevention of recurrent mood disorders. Pharmacopsychiatry. 2004 Nov;37(S 2):157–64.

53. Grahn JA, Parkinson JA, Owen AM. The cognitive functions of the caudate nucleus. Prog Neurobiol. 2008 Nov;86(3):141–55.

54. Shad MU, Muddasani S, Rao U. Gray matter differences between healthy and depressed adolescents: a voxel-based morphometry study. J Child Adolesc Psychopharmacol. 2012 Jun;22(3):190–7.

55. Kim MJ, Hamilton JP, Gotlib IH. Reduced caudate gray matter volume in women with major depressive disorder. Psychiatry Res. 2008 Nov 30;164(2):114–22.

56. Salgado S, Kaplitt MG. The nucleus accumbens: a comprehensive review. Stereotact Funct Neurosurg. 2015;93(2):75–93.

57. Munro CA, McCaul ME, Wong DF, Oswald LM, Zhou Y, Brasic J, et al. Sex differences in striatal dopamine release in healthy adults. Biol Psychiatry. 2006 May 15;59(10):966–74.

58. Belujon P, Grace AA. Dopamine system dysregulation in major depressive disorders. Int J Neuropsychopharmacol. 2017 Jun 29;20(12):1036–46.

59. Amico F, Meisenzahl E, Koutsouleris N, Reiser M, Möller HJ, Frodl T. Structural MRI correlates for vulnerability and resilience to major depressive disorder. J Psychiatry Neurosci. 2011 Jan 1;36(1):15–22.

60. Chen MC, Hamilton JP, Gotlib IH. Decreased hippocampal volume in healthy girls at risk of depression. Arch Gen Psychiatry. 2010 Mar 1;67(3):270–6.

61. Pagliaccio D, Alqueza KL, Marsh R, Auerbach RP. Brain volume abnormalities in youth at high risk for depression: adolescent brain and cognitive development study. J Am Acad Child Adolesc Psychiatry. 2020 Oct;59(10):1178–88.

62. Vogt BA, Palomero-Gallagher N. Chapter 25 - cingulate cortex. In: Mai JK, Paxinos G, editors. The Human Nervous System (Third Edition) [Internet]. San Diego: Academic Press; 2012 [cited 2023 Apr 6]. p. 943–87. Available from: https://www.sciencedirect.com/science/article/pii/B9780123742360100252

63. Berman MG, Peltier S, Nee DE, Kross E, Deldin PJ, Jonides J. Depression, rumination and the default network. Soc Cogn Affect Neurosci. 2011 Oct;6(5):548–55.

64. Koelsch S, Andrews-Hanna JR, Skouras S. Tormenting thoughts: The posterior cingulate sulcus of the default mode network regulates valence of thoughts and activity in the brain’s pain network during music listening. Hum Brain Mapp. 2021 Oct 15;43(2):773–86.

65. Li Q, Zhao Y, Chen Z, Long J, Dai J, Huang X, et al. Meta-analysis of cortical thickness abnormalities in medication-free patients with major depressive disorder. Neuropsychopharmacology. 2020 Mar;45(4):703–12.

66. Ducharme S, Albaugh MD, Nguyen TV, Hudziak JJ, Mateos-Pérez JM, Labbe A, et al. Trajectories of cortical thickness maturation in normal brain development--The importance of quality control procedures. NeuroImage. 2016 Jan 15;125:267–79.

67. Huttenlocher PR. Synaptic density in human frontal cortex - developmental changes and effects of aging. Brain Res. 1979 Mar 16;163(2):195–205.

68. Natu VS, Gomez J, Barnett M, Jeska B, Kirilina E, Jaeger C, et al. Apparent thinning of human visual cortex during childhood is associated with myelination. Proc Natl Acad Sci. 2019 Oct 8;116(41):20750–9.

69. Cowan WM. The development of the brain. Sci Am. 1979 Sep;241(3):113–33.

70. Maes M, Carvalho AF. The compensatory immune-regulatory reflex system (Cirs) in depression and bipolar disorder. Mol Neurobiol. 2018 Dec 1;55(12):8885–903.

71. Gegenhuber B, Tollkuhn J. Signatures of sex: Sex differences in gene expression in the vertebrate brain. WIREs Dev Biol [Internet]. 2020 Jan [cited 2023 Apr 6];9(1). Available from: https://onlinelibrary.wiley.com/doi/10.1002/wdev.348

72. McCarthy MM, Nugent BM, Lenz KM. Neuroimmunology and neuroepigenetics in the establishment of sex differences in the brain. Nat Rev Neurosci. 2017 Aug;18(8):471– 84.

73. Xu X, Coats JK, Yang CF, Wang A, Ahmed OM, Alvarado M, et al. Modular genetic control of sexually dimorphic behaviors. Cell. 2012 Feb 3;148(3):596–607.

74. Afroz S, Parato J, Shen H, Smith SS. Synaptic pruning in the female hippocampus is triggered at puberty by extrasynaptic GABAA receptors on dendritic spines. eLife. 2016 May 2;5:e15106.

75. Piekarski DJ, Boivin JR, Wilbrecht L. Ovarian hormones organize the maturation of inhibitory neurotransmission in the frontal cortex at puberty onset in female mice. Curr Biol. 2017 Jun 19;27(12):1735–1745.e3.

76. Wang Z, Gerstein M, Snyder M. RNA-Seq: a revolutionary tool for transcriptomics. Nat Rev Genet. 2009 Jan;10(1):57–63.

